# Predicting graft and patient outcomes following kidney transplantation using interpretable machine learning models

**DOI:** 10.1101/2023.08.24.23294535

**Authors:** Achille Salaün, Simon Knight, Laura Wingfield, Tingting Zhu

**Affiliations:** Institute of Biomedical Engineering, Department of Engineering, University of Oxford, Oxford, OX3 7DQ, United-Kingdom; Nuffield Department of Surgical Sciences, University of Oxford, Oxford, OX3 9DU, United-Kingdom

## Abstract

The decision to accept a deceased donor organ offer for transplant, or wait for something potentially better in the future, can be challenging. Clinical decision support tools predicting transplant outcomes are lacking. This project uses interpretable methods to predict both graft failure and patient death using data from previously accepted kidney transplant offers. Using more than twenty years of transplant outcome data, we train and compare several survival analysis models in single risk settings. In addition, we use *post hoc* interpretability techniques to clinically validate these models. Neural networks show comparable performance to the Cox proportional hazard model, with concordance of 0.63 and 0.79 for prediction of graft failure and patient death, respectively. Recipient and donor ages, primary renal disease, the number of mismatches at DR locus, and calculated reaction frequency at transplant appear to be important features for transplant outcome prediction. Owing to their good predictive performance and the clinical relevance of their *post hoc* interpretation, neural networks represent a promising core component in the construction of future decision support systems for transplant offering.

## Introduction

Around 2,500 deceased donor kidney transplants are performed in the UK each year. At any time, there are around 5,000 patients on the kidney transplant waiting list with an average wait of 2-3 years. The shortage of organs available for transplant means that some patients become unfit for surgery or die whilst waiting. Because of this, clinicians often consider organ offers from less-than-optimal donors with existing comorbidities or older age. Decisions around organ offers are made by clinicians based upon the information available at the time of offer, including donor and recipient demographic and medical details. Clinicians use their clinical experience, but do not have reliable tools available to help them predict what would happen if they choose to accept or decline an offer and wait for the next available one. This uncertainty leads to considerable variability in organ decline rates and waiting times between clinicians and centres. A clinical decision support (CDS) system that accurately predicts transplant outcomes, both in terms of graft failure and patient death, as well as indicating what would happen if the organ offer was declined (in terms of future offers and likely waiting time), may help to support clinicians in making these difficult decisions. As decisions must remain under the responsibility and control of the clinician, any CDS tool must be easy to use, and predictions must be interpretable from a clinician’s perspective. Interpretability and usability are also important to patients, allowing better explanations of likely outcomes during the informed consent process.

The aim of this study is to predict transplant outcomes in the scenario of an accepted kidney offer. We utilise more than twenty years of registry data, containing over 36,000 accepted kidney transplant offers, with graft and patient survival information. These data have been provided from the National Health Service Blood and Transplant (NHSBT) UK Transplant Registry with ethical approval. Using these data, we have trained and compared several survival analysis models. In addition, we use *post hoc* interpretability techniques to clinically validate these models.

Predicting the time of occurrence of an event (such as patient death or graft failure) from censored data has been extensively studied under the name of survival analysis. This has many applications in health informatics such as predicting strokes [1], oral cancer [2], or graft outcome prediction. Censored data are common in such contexts, resulting from loss of follow-up, competing events, or the end of the study. In the context of graft outcome prediction, previous studies use the Cox proportional hazard (PH) model to predict kidney graft or recipient survival [3–5]. The Cox PH model is a classic time-to-event approach that models the hazard function, as in the failure rate of a system according to time [6]. This approach is not only robust and reliable, but also simple to use and well understood by clinicians. Several generalisations of this model have been proposed. For instance, DeepSurv [7] aims at increasing the modelling power of the Cox model by replacing the linear contribution of the covariates with a neural network. DeepHit [8] directly models the cumulative incidence function with a single neural network. Originally proposed for handling competing risks, this network is structured according to a multi-task architecture, composed of a shared sub-network and several cause-specific sub-networks. The loss has been adapted to maximise the concordance index, a classic survival analysis metric based on the idea that the earlier an event is observed, the higher the associated risk should be.

In contrast to the Cox PH model, this approach does not rely on the proportional hazard assumption. Neural networks are not the only machine learning models that have been adapted to survival analysis. For instance, random survival forests [9] is an adaptation of random forests to right-censored survival data. Alternatively, classification machine learning methods can be considered to predict the status of the subject of interest at specific time points. For example, predicting transplant outcomes after 1, 5, and 10 years is generally sufficient for the clinicians. Thus, existing risk communication tools such as [10] identify survival functions obtained from the Cox PH model at these time points. Many previous publications follow this classification approach [3, 11, 12]. However, this approach requires to train a model for each time point. These independent models may induce inconsistent results when considered all together. Whilst many of these previous studies demonstrate acceptable predictive performance, none challenged their models’ validity through the lens of clinical interpretability.

Interpretability is another important criterion in the construction of a CDS tool for predicting graft outcomes. Interpretability is the extent to which the prediction of a model can be understood by a human [13]. This way, users can build trust regarding the model’s results and remain in control of the associated outcomes. A good model should always be *intrinsically* interpretable to a certain degree. Indeed, interpretable models have been shown to be more robust to adversarial attacks [14]. Although some of the approaches mentioned above, like the Cox PH model, are inherently interpretable, some other models, like neural networks, are designed in a way that makes interpretation difficult. Nonetheless, it is possible to interpret *a posteriori* a black box neural network model with the help of *post hoc* interpretability methods. One can provide a local explanation of a given prediction. For instance, LIME [15] locally samples data points around the input and returns a linear explanation of the predictions made by the black-box model from these data points. Unfortunately, this solution is unstable; explanations depend highly on the sampled data points, harming the trustworthiness of the explanations. Similarly to LIME, SHAP [16] is a *post hoc* interpretability method relying on additive feature attribution models, i.e. linear functions as local explanation models. It provides explanations *via* game theory: each prediction is seen as a game where the features are players contributing to that game. Feature contributions are computed by considering all possible coalitions of features and the marginal contribution of each feature within these coalitions. Hence, SHAP can be considered as a gold standard in terms of *post hoc* interpretability methods.

## Methods

All methods were carried out in accordance with relevant guidelines and regulations. This study, referenced under IRAS project ID 304542, has received approval from the Health Research Authority and Health and Care Research Wales (UK research ethics committee).

### Data

Our work is based on the analysis of a data set from the UK Transplant Registry, provided by NHSBT. It describes 36,653 accepted kidney transplants, performed between the years 2000 and 2020, across 24 UK transplant centres. The total follow-up duration is around 22 years. Each transplant is originally described with 3 identifiers, 12 immunosuppression follow-up indicators, 143 donor, recipient and transplant characteristics, and 7 entries describing targeted outcomes. Considering transplants as independent, we exclude the transplant, donor, and recipient identifiers. Information regarding post-transplant immunosuppression is discarded as this is not available at the time of the offer decision. The donor, recipient and transplant characteristics serve as input features for modelling. Among them, 24 describe the recipient, 109 represent the donor, and 10 refer to the overall transplant. Both recipient and donor characteristics contain generic information such as gender, ethnicity, age, blood group, height, weight, or body mass index (BMI). More specific information is also available, such as the transplant centre, number of previous transplants, waiting time, ease of matching, and the dialysis status. Donor data include the cause of death, past medical history and results of blood tests including kidney function (estimated glomerular filtration rate, eGFR). Transplant data include the donor-recipient immunological match.

Duplicate rows are removed, and values outside of a plausible clinical range are removed. Categorical values are checked by clinicians and simplified (or removed) if needed. BMI is recomputed based on weight and height. Both weights and heights are discarded to limit redundant information. Blood measurements are harmonised across the data set by selecting the first measurement available (generally at donor registration) and the maximum value during the donation process. Since the calculation of eGFR varies across hospitals, this metric is recomputed over the whole data set using a consistent definition (see appendix, section A.1). Recipient dialysis status is also simplified into a dialysis duration and dialysis modality at time of transplant (predialysis, haemodialysis or peritoneal dialysis). Transplant offers not meeting the inclusion criteria, such as dual and multi-organ transplants, are discarded.

Outcomes present in the dataset include information about graft failure, patient death, and transplant failure. Graft failure excludes death with a functioning graft, whilst transplant failure denotes either graft failure or death. In this work, we focus on predicting graft failure and patient death. Each outcome is represented as a pair containing an event time and a right-censoring indicator. Right-censoring is a common type of censoring in survival analysis that describes the loss of follow-up on the event of interest. It can occur for various reasons, such as the end of the study, competing events, etc. Thus, right-censored information provides some partial information about the survival time, where it is only known to be greater than the censoring time. Transplant outcomes are recomputed for the sake of consistency.

After removing the features presenting more than 50% missing values across the whole data set, the data is described through 50 input variables. At this stage, the data contains 8% missingness. A summary of this data-cleaning process is given in Figure 1. Additionally, an exhaustive list of the features and targets considered at the latest stage of this process is given in the appendix (section A.2).

**Figure 1.**
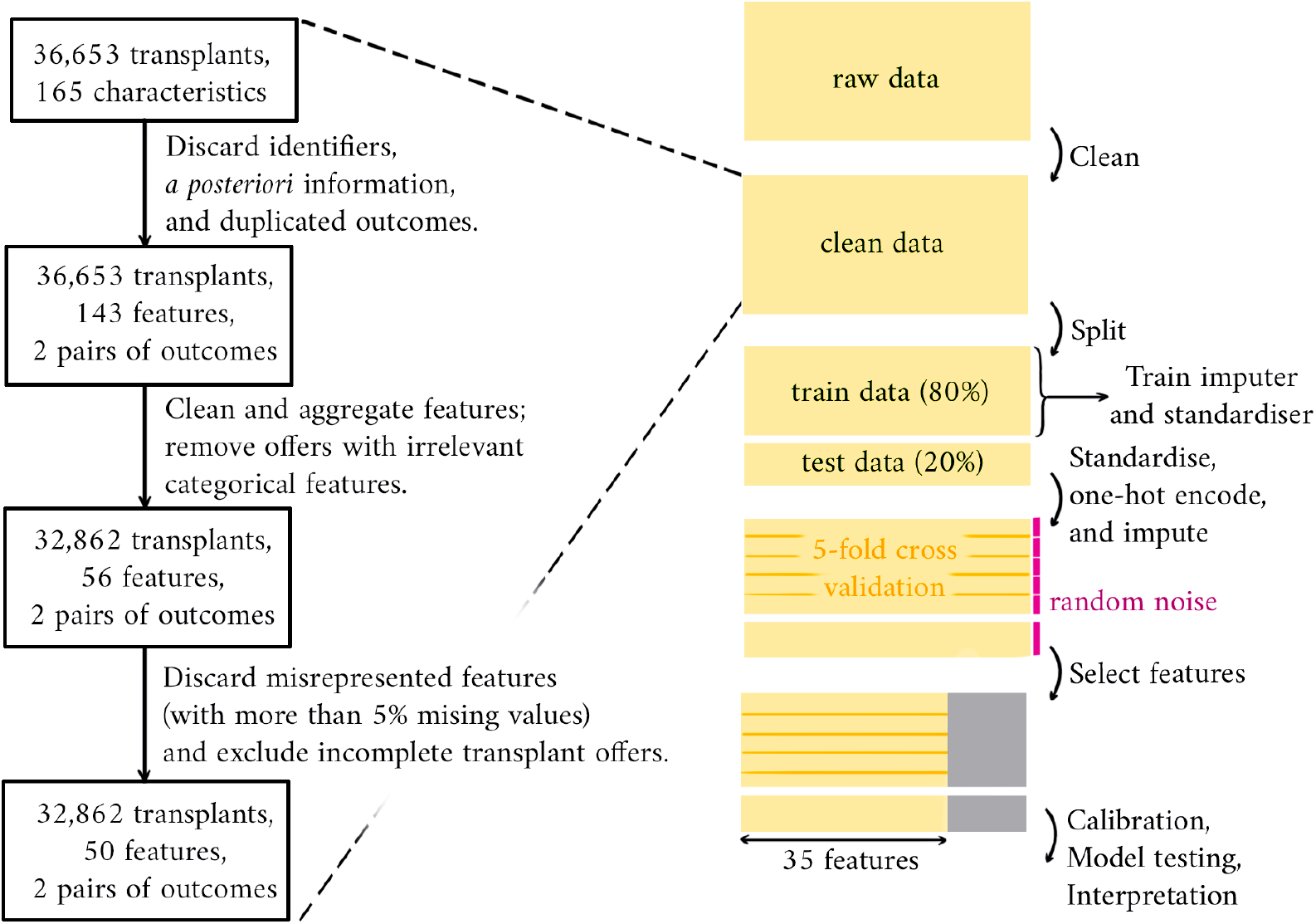
End-to-end data processing pipeline, from raw data to model testing. Data cleaning is detailed on the left. Cross-validation is performed before and after feature selection.

### Model Training and Validation

In this article, we compare the Cox PH model, DeepHit, and random survival forests in a single risk setting. The different models are interpreted *a posteriori*, and their performances are discussed.

The following methodology is applied. First, the data is split in a stratified manner with regards to censoring indicators, where 80% of the data is reserved for training and the remaining 20% is left for testing. Due to matching policy changes and follow-up time differences between old and recent offers, we do not split the data according to transplant dates. After this first step, numerical values are standardised and categorical ones are one-hot-encoded. Mean and variance are computed over training data only. Standardisation has appeared to be more relevant than normalisation due to the presence of outliers in the data. Then, we impute missing features with the help of MissForest, an iterative imputation technique relying on random forests [17]. MissForest is first trained on the training data and then applied to all the data. This solution has been selected among several imputation techniques including MIDAS, a variational autoencoder-based imputation technique [18]; MICE, an iterative method for multi-column imputation [19]; MissForest itself, which is a variant of MICE; and a naive imputer simply returning average values. These methods have been compared on a sample of the data, where missingness was introduced by randomly masking known values. After this imputation step, survival analysis models are trained through 5-cross validation. This process is performed a first time for feature selection. This is achieved by inputting Gaussian noise as a feature: we select any feature whose importance is higher than the importance attached to noise. Based on this subset of features, 5-cross validation is then repeated for final model training. Model calibration is then performed: predictions are adjusted *a posteriori* with the help of a logistic function to match observed outcomes ratios. Model evaluation is undertaken by computing concordance and AUROC scores over 100 bootstraps of the testing data. The survival models are clinically interpreted using SHAP. To do so, we fix a particular time point (1, 5, or 10 years) and consider how models predict event occurence up to that point. The coefficients of the Cox PH model are also provided. The Figure 1 illustrates the overall methodology and the Python code used for experiments can be found at https://github.com/AchilleSalaun/Xamelot.

Following this processing pipeline, we compare the Cox PH model, random survival forests, and neural networks. Since both DeepHit and survival forests require the time to be discretised, we restrict transplant outcome prediction to 1, 5, and 10 years. This step follows the discretisation process described in [8]. We rely on grid search to tune hyperparameters. As a result, Breslow’s estimator is used to derive the Cox PH model’s baseline [20]. In addition, a regularisation parameter is introduced and set to 1*e*^−4^ to deal with colinearities in the data. The survival random forest is given 300 trees. Finally, we train DeepHit in a single risk fashion. While predicting graft failure, the model is instantiated with two hidden layers of 100 neurons, with 10% dropout. The neural network used to predict patient death shows one hidden layer of 200 neurons followed by two layers of 100 neurons. For both graft failure and patient death predictions, the training is done through 50 epochs, with batches of size 64, and a learning rate equal to 1*e*^−2^.

## Results

### Single Outcome Prediction

In total, 35 features are selected to predict graft failure and patient death. Recipient and donor ages, primary renal disease, the number of mismatches at DR locus, and calculated reaction frequency at transplant are important features for prediction of both outcomes.

Table 1 displays the concordance scores reached by each model, for both graft failure and patient death prediction. Tables 2 and 3 provide the AUROC reached by each model on predicting graft failure and patient death, respectively, for observations years 1, 5, and 10. Performances before and after feature selection are presented. One can observe that overall performances increase with the observation time, being maximal at year 10. This may be explained by the fact that the features present in the original dataset are more relevant to long term predictions than short term ones. Also, event rates increase over time. Considering both concordance and AUROC, the neural network shows similar performances to the performances of the random forest and the Cox PH model, slightly outperforming them on the graft failure prediction task.

**Table 1.** Concordance scores for graft failure and patient death prediction. Best results are highlighted in bold.

|  |  | Random forest | Cox PH model | Neural network |
| --- | --- | --- | --- | --- |
| Graft failure | Before feature selection | .623 ($\pm 1e^{-4}$) | .610 ($\pm 1e^{-4}$) | **.625** ($\pm 1e^{-5}$) |
| | After feature selection | .626 ($\pm 1e^{-4}$) | .612 ($\pm 1e^{-4}$) | **.630** ($\pm 1e^{-4}$) |
| Patient death | Before feature selection | .782 ($\pm 1e^{-4}$) | **.792** ($\pm 1e^{-4}$) | .791 ($\pm 1e^{-4}$) |
| | After feature selection | .788 ($\pm 1e^{-4}$) | **.792** ($\pm 1e^{-4}$) | .790 ($\pm 1e^{-4}$) |

**Table 2.** AUROC with respect to graft failure prediction. Best scores are highlighted in bold.

|  |  | Random forest | Cox PH model | Neural network |
| --- | --- | --- | --- | --- |
| Year 1 | Before feature selection | .620 ($\pm 2e^{-4}$) | .606 ($\pm 2e^{-4}$) | **.621** ($\pm 2e^{-4}$) |
| | After feature selection | .614 ($\pm 2e^{-4}$) | .603 ($\pm 2e^{-4}$) | **.626** ($\pm 2e^{-4}$) |
| Year 5 | Before feature selection | **.641** ($\pm 1e^{-4}$) | .626 ($\pm 1e^{-4}$) | .616 ($\pm 1e^{-4}$) |
| | After feature selection | **.646** ($\pm 1e^{-4}$) | .630 ($\pm 1e^{-4}$) | .629 ($\pm 1e^{-4}$) |
| Year 10 | Before feature selection | .670 ($\pm 1e^{-4}$) | .657 ($\pm 1e^{-4}$) | **.676** ($\pm 1e^{-4}$) |
| | After feature selection | .669 ($\pm 1e^{-4}$) | .662 ($\pm 1e^{-4}$) | **.686** ($\pm 1e^{-4}$) |

**Table 3.** AUROC with respect to patient death prediction. Best scores are highlighted in bold.

|  |  | Random forest | Cox PH model | Neural network |
| --- | --- | --- | --- | --- |
| Year 1 | Before feature selection | .739 ($\pm 3e^{-4}$) | **.749** ($\pm 3e^{-4}$) | .747 ($\pm 3e^{-4}$) |
| | After feature selection | .734 ($\pm 3e^{-4}$) | **.747** ($\pm 3e^{-4}$) | .744 ($\pm 3e^{-4}$) |
| Year 5 | Before feature selection | .793 ($\pm 1e^{-4}$) | **.804** ($\pm 1e^{-4}$) | .801 ($\pm 1e^{-4}$) |
| | After feature selection | .798 ($\pm 1e^{-4}$) | **.804** ($\pm 1e^{-4}$) | .802 ($\pm 1e^{-4}$) |
| Year 10 | Before feature selection | .833 ($\pm 1e^{-4}$) | **.845** ($\pm 1e^{-4}$) | .844 ($\pm 1e^{-4}$) |
| | After feature selection | .837 ($\pm 1e^{-4}$) | **.845** ($\pm 1e^{-4}$) | .842 ($\pm 1e^{-4}$) |

From an interpretability viewpoint, the neural network, when combined with SHAP, provides a richer clinical depiction of the data than Cox. The features that are important to clinicians are also considered important to the neural network. For example, among predominant features for graft failure prediction (cf. Figures 2.a and 2.b), recipient and donor age, donor type, donor past hypertension, or eGFRs are also features commonly used by regression models from the transplant literature [4, 21, 22]. The direction of effect of feature values on predictions also matches clinical knowledge. For instance, patients with diabetes are likely to have inferior survival. This is reflected through the higher SHAP values regarding graft failure when `prd#Diabetes` is equal to one. The effect of covariates on survivability can be non-linear, as illustrated by the recipient age (`rage`; see Figure 2.c). Indeed, it is commonly recognised that younger patients can be less adherent to medication, hence increasing the risk of graft failure. This phenomenon vanishes with older patients, and age then becomes a penalising feature for survivability. In contrast, explanations obtained from the Cox PH model do not highlight such behaviours (see Figure 2.d), being limited to less expressive covariate effects. By design, it can be summarised as a linear function in the case of Cox. Moreover, Cox coefficients do not seem to reflect clinical expertise in terms of feature importance. For example, it attaches a strong importance to the hospital centre while both neural networks and random forests agree on the predominance of donor age. Finally, survival random forests share similar interpretations to DeepHit.

**Figure 2.**
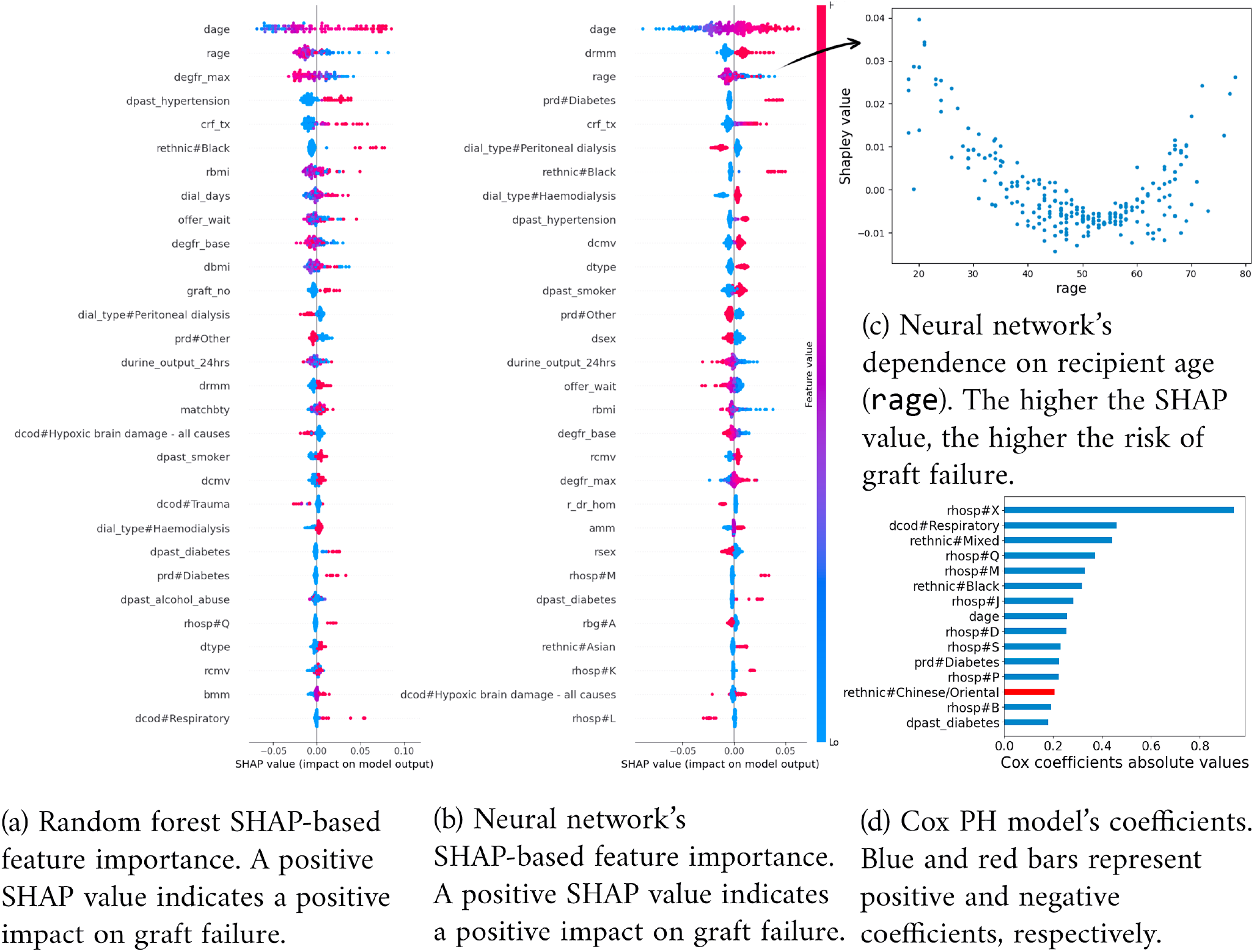
Interpreting graft failure prediction at 10 years. Sub-figures (a) and (b) provide covariate effects with regards to SHAP for random forests and neural networks, respectively. Each point represents a Shapley value for a particular feature in a particular offer. The values of the features are represented through colour: blue and pink indicate features whose values are low and high, respectively. When focusing on a single feature, this information can be directly reported onto the y-axis (cf. sub-figure (c)).

## Discussion

Neural networks have shown comparable performance to tools generally used by clinicians when predicting kidney transplant outcomes. In particular, they perform well when predicting long-term outcomes, which is a useful property when making a decision as to whether to accept an organ offer.

The Cox PH model remains a robust solution in terms of performance, with little to no hyperparameter tuning. It is simple to use, leads to reliable predictions, and is easy to understand by clinicians. However, whilst Cox PH models can be interpreted at a model level by inspecting regression coefficients, interpretability at an individual prediction level is not as easy. First, feature effects are linked indirectly to the survival function through the linear component in the hazard function. Since Cox PH model’s inherent interpretability comes from this linear component, variants aiming at modelling non-linear covariates (e.g. by use of splines) might hamper interpretability. Furthermore, using splines would require an *a priori* understanding of the covariate effects. Finally, interpretations do not depend on the prediction time due to the proportional hazard assumption.

In contrast, and despite their black-box nature, neural networks stand out in terms of interpretability. Using SHAP allows us to have fine-grain, real-time interpretations for individual predictions, which is not possible with the Cox PH model. This level of interpretability allows us to clinically validate these models, making them more trustworthy and explainable to patients. SHAP can also highlight interesting relationships between covariates and transplant outcomes. Previous analyses of the UK registry data show that outcomes from kidney donations before and after circulatory death are equivalent regarding both patient and graft survival [23, 24]. Our models, trained from a larger dataset, suggest that donor type can have an impact on long-term transplant outcomes (see Figure 2.b, `dtype`). This level of interpretability is useful to explain individual prediction through the lens of SHAP. For example, Figure 4 shows the contribution of each feature to a given prediction of graft survival at year 10. In this case, predicted survivability is mainly lowered by the calculated reaction frequency at transplant and the donor type. Survival forests exhibit comparable performance and interpretation to neural networks. Nevertheless, the process of training and using random forests is considerably slower compared to training and using neural networks or the Cox PH model. Consequently, neural networks emerge as more preferable candidates when compared to survival forests.

**Figure 3.**
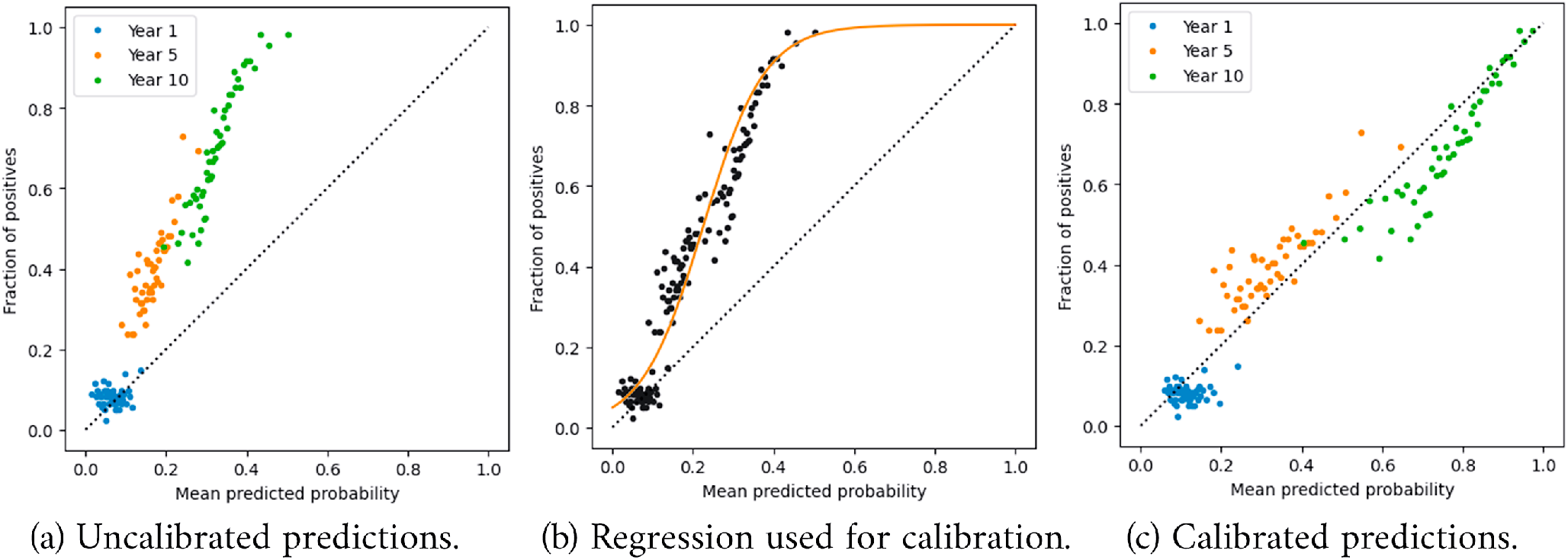
Calibration of the neural network predicting graft failure. Each point represents a cohort of transplant offers that share similar predicted scores: the average score respective to each group is reported on the x-axis and the true class ratio within each groups is reported on the y-axis. Predictions at 1, 5, and 10 years are considered all together while training the calibration model.

**Figure 4.**
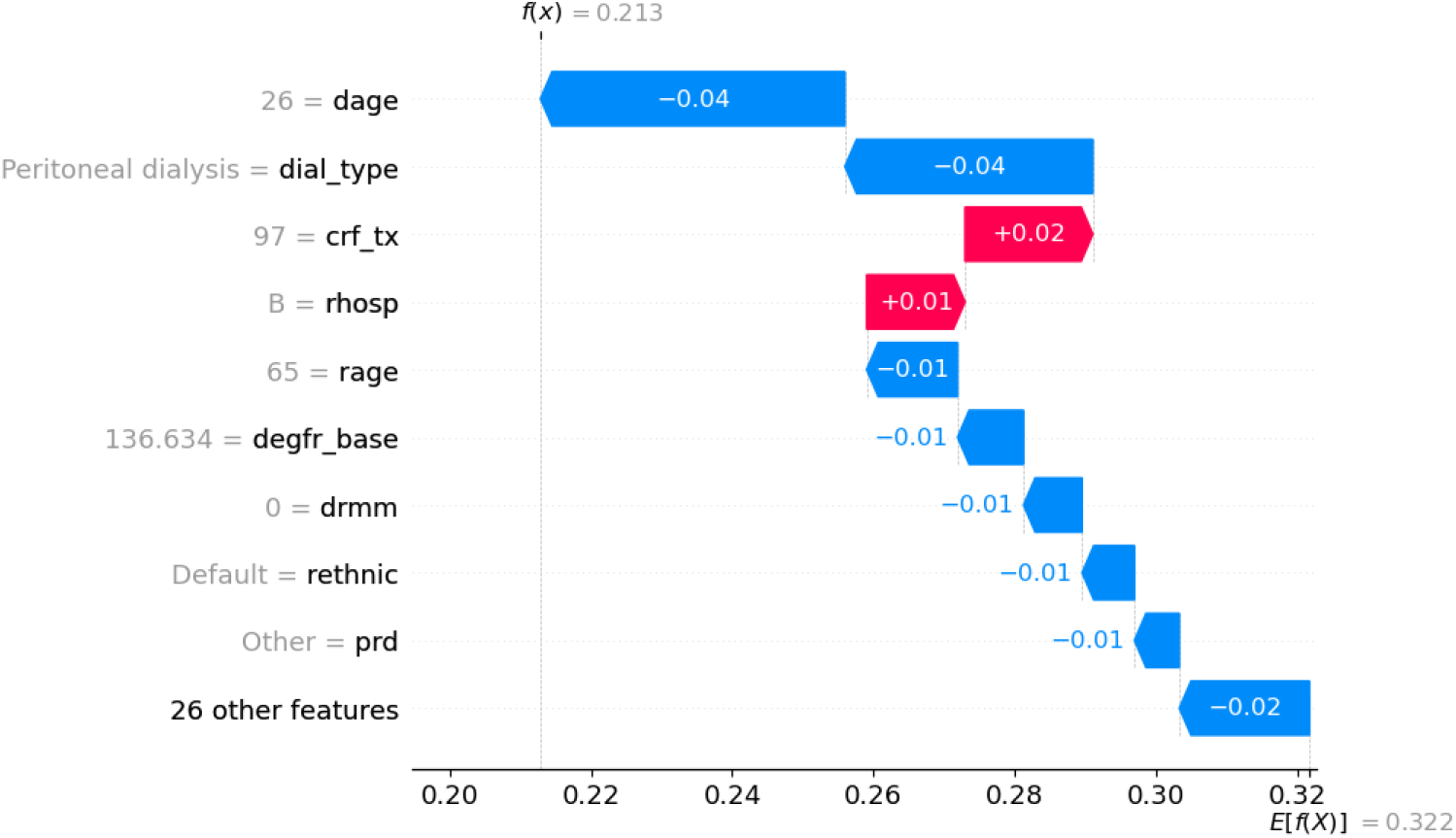
Waterfall plot for an example prediction of graft failure at 10 years. Default recipient ethnicity is white. Positive SHAP values indicate a positive impact on graft failure, and *vice versa*. The plot demonstrates the impact of individual features in moving the predicted survival away from the population average (0.322) to an individualised prediction (0.213).

As shown in Figure 3, calibration remains a crucial step in the process: as prediction scores are meant to support clinicians’ decisions, it is important to ensure that these scores offer an intuitive representation of the risks associated with graft failure and patient death. It is worth noting that calibration does not alter raw performances, provided the calibration function is strictly increasing. Indeed, concordance assesses how well data points are sorted, while AUROC evaluates how effectively they are classified. In this survival analysis context, considering predictions at years 1, 5, and 10 all together while performing calibration prevents introduction of inconsistencies in predictions (e.g predicting a higher probability of graft failure at year 5 than at year 10).

Let us now discuss the limitations and possible research directions regarding this work. First, the data reflects more long term outcomes than early ones. For instance, features like kidney anatomy and damages are missing. Including such features could improve performances at shorter terms. More importantly, predicting transplant outcomes is only one aspect of the construction of a CDS tool for kidney offering. However, predicting what could be the consequences of refusing an organ offer in terms of future transplant opportunities, death, or removal from the waiting list is another key step. Having a good understanding of the outcomes in both scenarios is indeed necessary to predict individualised treatment effects. In parallel, measures to safe-guard the use of the CDS tool are necessary. If interpretability contributes to building trust regarding the tool’s predictions, uncertainty quantification is another critical feature regarding the construction of a CDS tool for organ offering. This can be achieved either through post hoc error prediction using meta-modeling, or with a Bayesian version of our models. The presence of biases derived from the data is also something to inspect more carefully. As our models have been compared on a fair ground, possible biases should not impact our conclusions. Preliminary tests show optimistic results regarding the limitation of biases related to sensitive characteristics like age, gender, or ethnicity. One can also note that we do not use SHAP to explore inter-covariate dependencies: if it could be relevant in the context of the clinical model validation, such interactions start to be difficult to explain and present to clinicians in practice. Finally, we have yet to address the aspect of maintainability for our tool. This will require recurring validation on recent data as it becomes available, with retraining of the models if performance decreases. The optimum solution for this would be continuous learning, but this may be challenging in the context of healthcare data due to limitations in access to datasets.

To conclude, we have trained several models to predict transplant outcomes from kidney offers, based on twenty years of registry data. Neural networks provide comparable results to classic survival analysis models. By using SHAP, we provide clinically validated interpretations of these models. This level of interpretability is especially relevant to enable validation from clinicians and to involve patients in the decision-making process. Therefore, neural networks represent a promising core component in the construction of future CDS system for transplant offering. As future work, we want to extend our analysis to the prediction of patient outcomes in the case of a declined offer. We also plan to add uncertainty quantification to our CDS tool.

## Data availability statement

The dataset analysed during the current study is not publicly available due to the property of NHSBT but is available from the corresponding author on reasonable request.

## Acknowledgements

The authors thank the anonymous reviewers for their valuable suggestions. This work has been supported by funds from the NIHR (AI Award 2020 Phase 1: `AI_AWARD02316`). T.Z. was supported by the Royal Academy of Engineering under the Research Fellowship scheme.

## Author contributions statement

A.S. undertook the data cleaning, model building, and redaction of this paper. S.K. and L.W. provided clinical input to data cleaning and model design. S.K. and T.Z. co-ordinated the overall project, providing respectively clinical and machine learning insights.

## Additional information

No competing interest is declared.

