## Appendix for "Predicting graft and patient outcomes following kidney transplantation using interpretable machine learning models"

### A.1 Estimation of eGFR

Estimated glomerular filtration rate (eGFR) is a score that represents the kidney filtration rate and is derived from various measurements such as creatinine. It can be derived according to the following formulas. Adult (older than 16 years old) eGFR [1] is derived from the formula

$$\begin{aligned}\mathrm{eGFR_{adult}} &\stackrel{\text{def}}{=} 142 \\ &\times \min\left(\frac{\text{creatinine}}{0.7\delta_\mathrm{f} + 0.9(1-\delta_\mathrm{f})}, 1\right)^{-0.241\delta_\mathrm{f} - 0.302(1-\delta_\mathrm{f})} \\ &\times \max\left(\frac{\text{creatinine}}{0.7\delta_\mathrm{f} + 0.9(1-\delta_\mathrm{f})}, 1\right)^{-1200} \\ &\times 0.9938^{\text{age}} \times 1.012^{\delta_\mathrm{f}},\end{aligned}$$

where $\delta_f$ is equal to 1 if the patient is female, or 0 else. The creatinine measurements need to be expressed in mg/dl and the provided measurements be expressed in $\mu$mol/l, the latter is first converted as follows:

$$\text{creatinine}_{\text{mg/dl}} \stackrel{\text{def}}{=} 113.1222\text{e}{-4} \times \text{creatinine}_{\mu\text{mol/l}}.$$

Pediatric eGFR [2] is given by

$$\mathrm{eGFR_{pediatric}} \stackrel{\text{def}}{=} 0.413 \times \frac{\text{height}}{\text{creatinine}},$$

where creatinine is also in mg / dl, and height in cm.

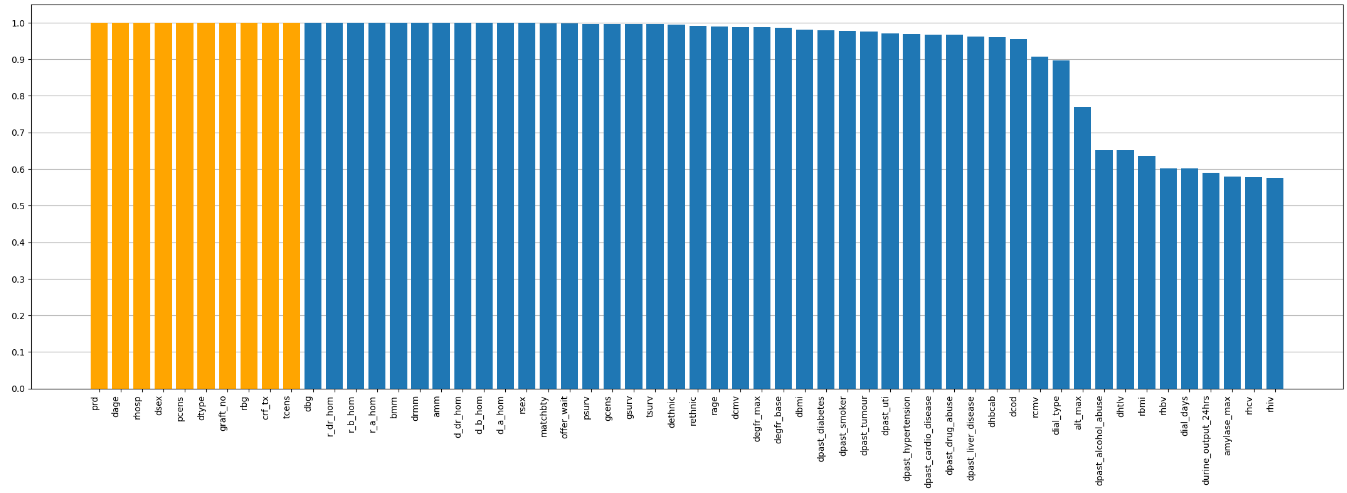


Figure 1: Features densities. A density of 1 means that the corresponding feature is always present; conversely, a density of 0 means that this feature is always missing. Orange columns highlight features with no missing value at all.

### A.2 Features

Figure 1 shows the proportion of non-missing values for each feature in the data after cleaning. Features with a density lower than 0.5 are not displayed.

The following is a statistical overview of the data after cleaning. For each feature, this overview includes a display of the feature's distribution and its density. If a feature is numerical, we also provide its minimum, maximum, mean values, and its standard deviation. When a categorical feature is binary, we include the mapping between the labels and $\{0, 1\}$. We limit this overview to the 35 features resulting from the feature selection process (see section "Methods", subsection "Model Training and Validation"). Similarly, we also add a description of the targets at the end of this overview.

To enhance readability, each feature and target is presented on its own page.

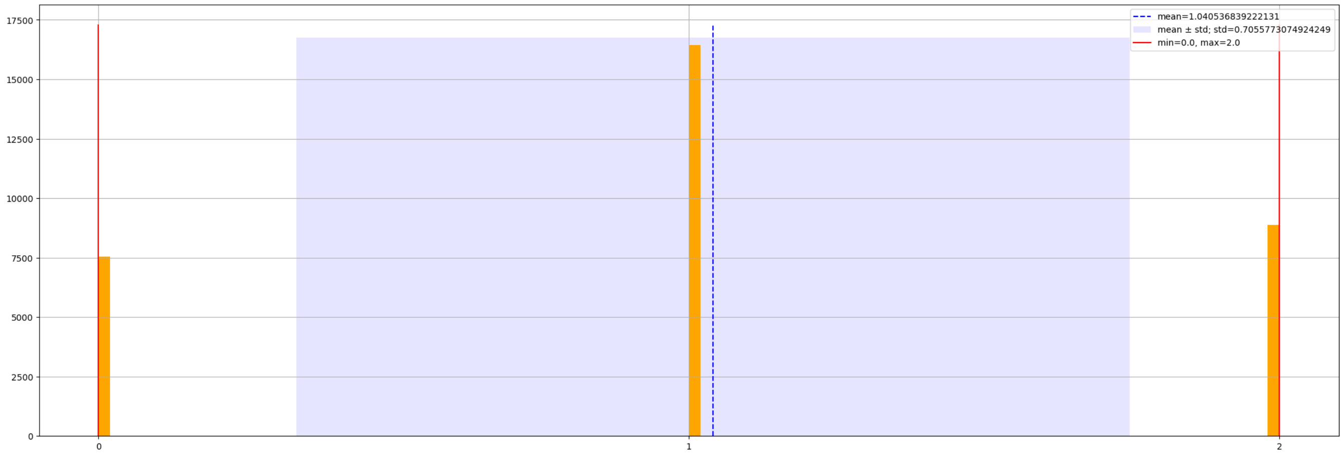


Figure 2: Feature distribution: amm.

**amm**

**Description:** Number of mismatches at A locus.

**Type:** Numerical

**Density:** 1.0

**Mininimum:** 0.0

**Maximum:** 2.0

**Mean:** 1.0

**Standard deviation:** 0.7

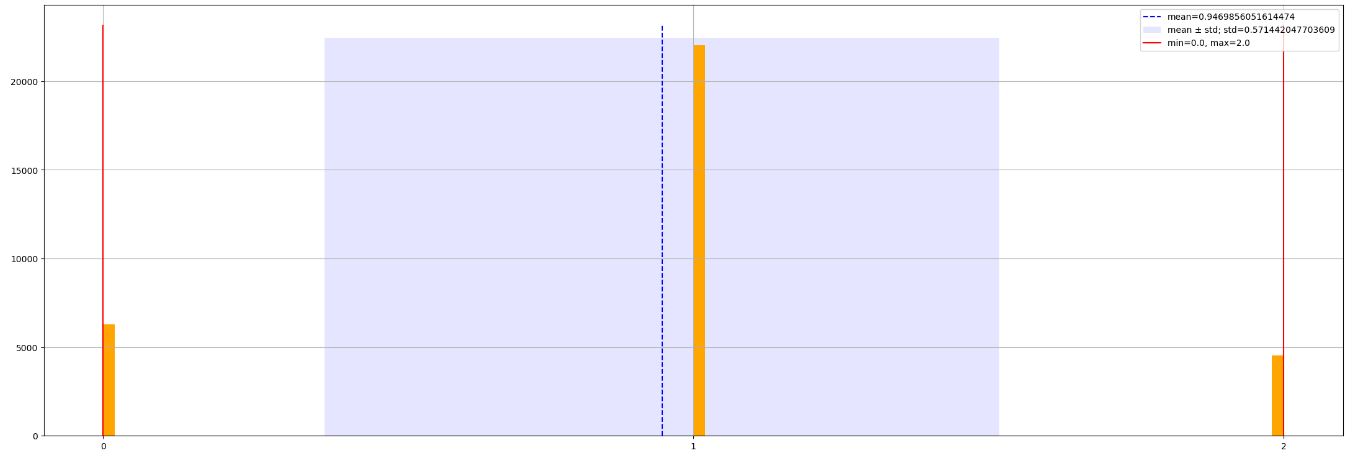


Figure 3: Feature distribution: bmm.

**bmm**

**Description:** Number of mismatches at B locus.

**Type:** Numerical

**Density:** 1.0

**Mininimum:** 0.0

**Maximum:** 2.0

**Mean:** 1.0

**Standard deviation:** 0.6

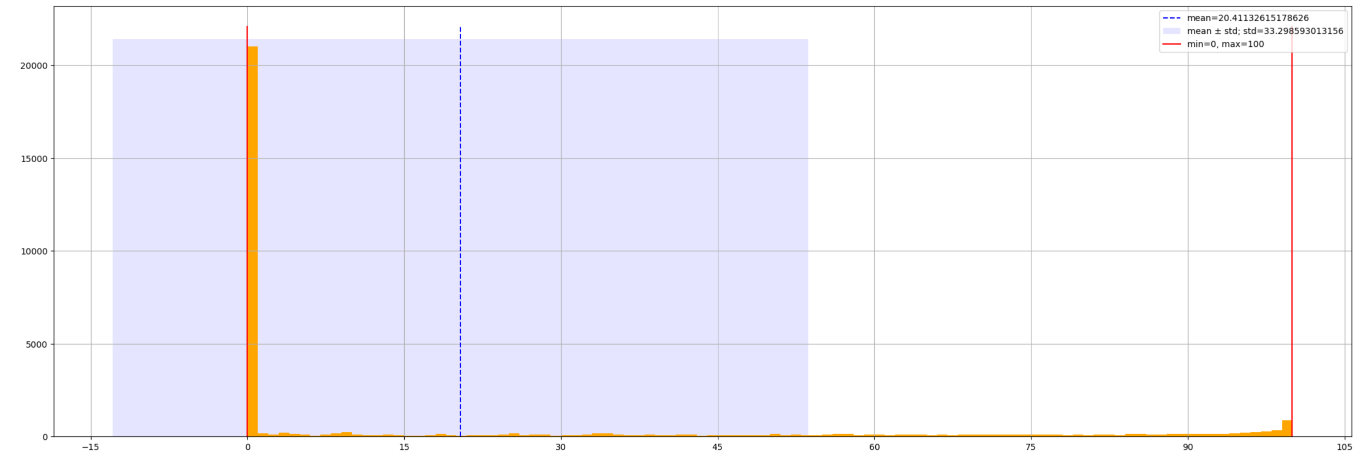


Figure 4: Feature distribution: crf_tx.

**crf_tx**

**Description:** Calculated reaction frequency at transplant.

**Type:** Numerical

**Density:** 1.0

**Mininimum:** 0.0

**Maximum:** 100.0

**Mean:** 20.4

**Standard deviation:** 33.3

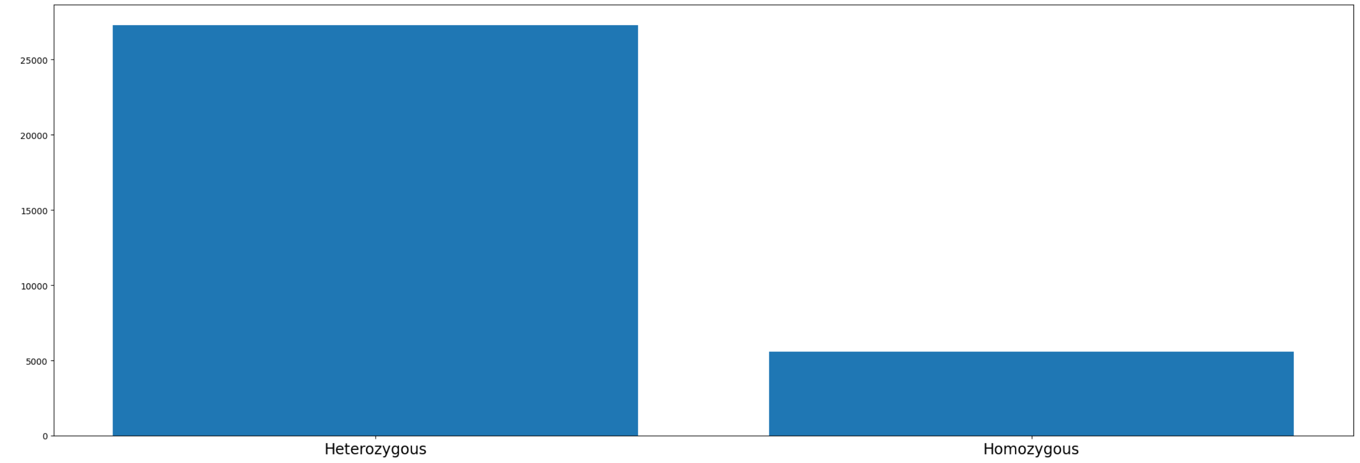


Figure 5: Feature distribution: d_a_hom.

### d_a_hom

**Description:** Donor homozygous at A locus.

**Type:** Categorical

- Heterozygous : 0
- Homozygous : 1

**Density:** 1.0

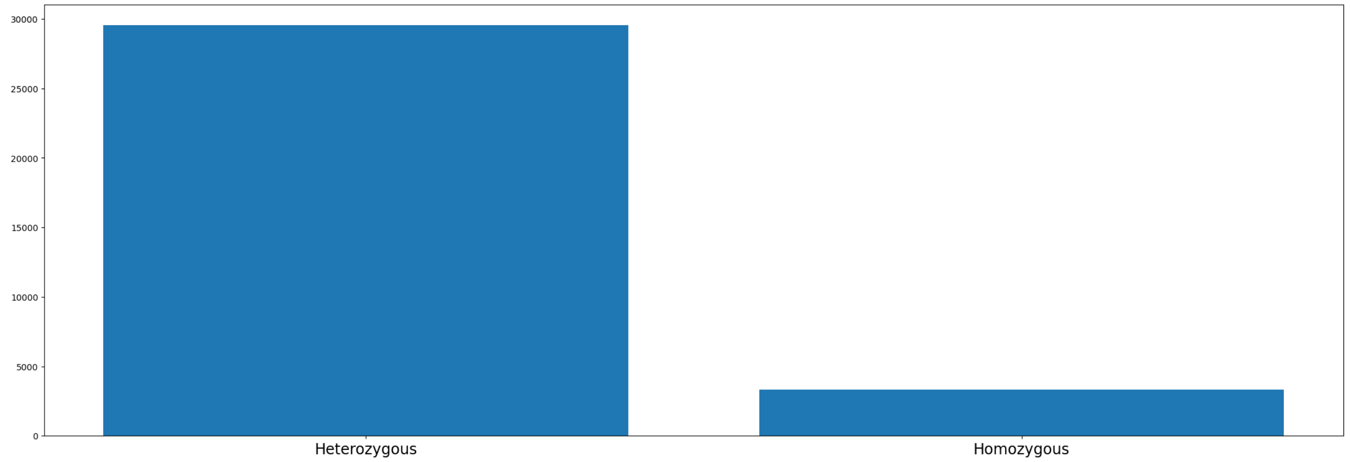

Figure 6: Feature distribution: d_b_hom.

#### d_b_hom

**Description:** Donor homozygous at B locus.

**Type:** Categorical

- Heterozygous : 0
- Homozygous : 1

**Density:** 1.0

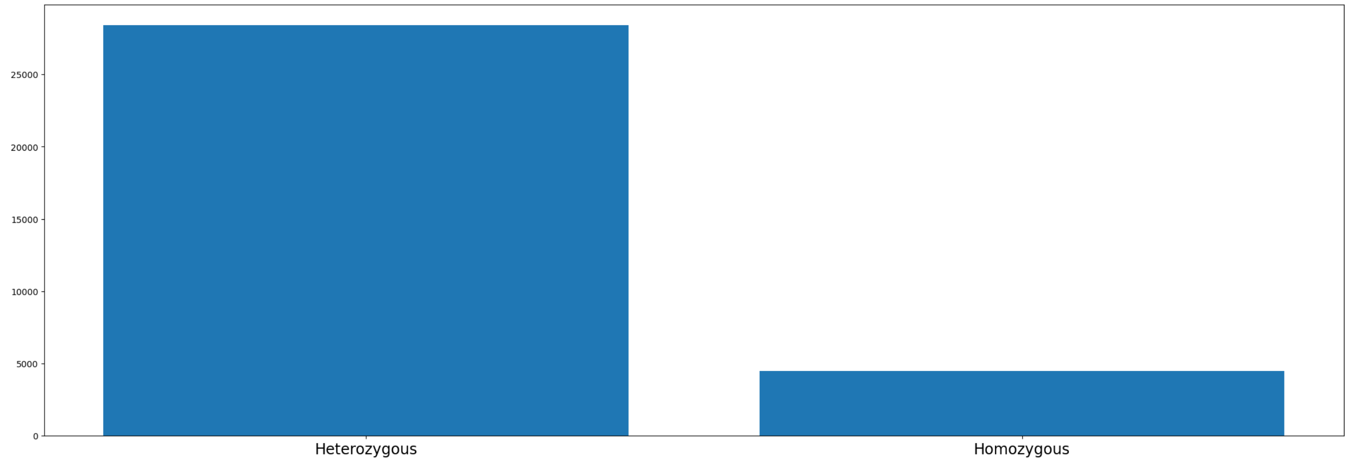


Figure 7: Feature distribution: d_dr_hom.

**d_dr_hom**

**Description:** Donor homozygous at DR locus.

**Type:** Categorical

- Heterozygous : 0
- Homozygous : 1

**Density:** 1.0

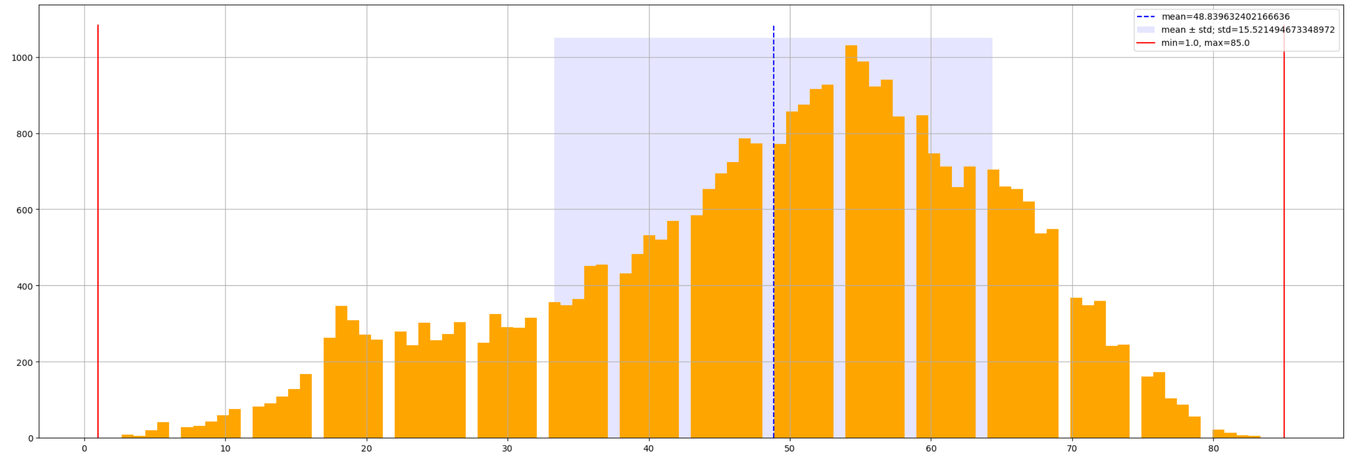


Figure 8: Feature distribution: dage.

#### dage

**Description:** Donor age.

**Type:** Numerical

**Density:** 1.0

**Mininimum:** 1.0

**Maximum:** 85.0

**Mean:** 48.8

**Standard deviation:** 15.5

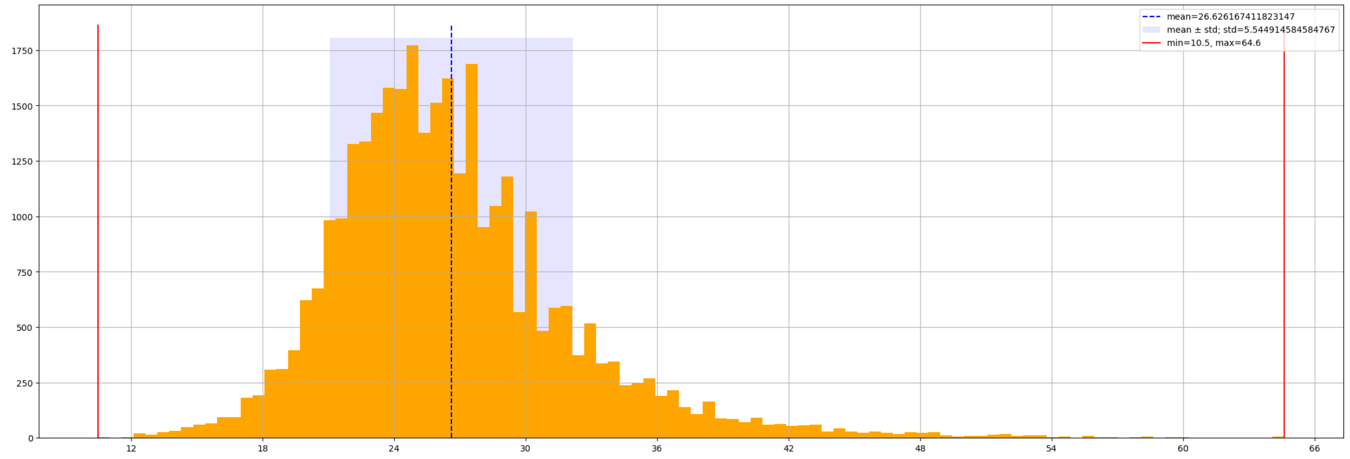


Figure 9: Feature distribution: dbmi.

#### dbmi

**Description:** Donor BMI.

**Type:** Numerical

**Density:** 1.0

**Mininimum:** 10.5

**Maximum:** 64.6

**Mean:** 26.6

**Standard deviation:** 5.6

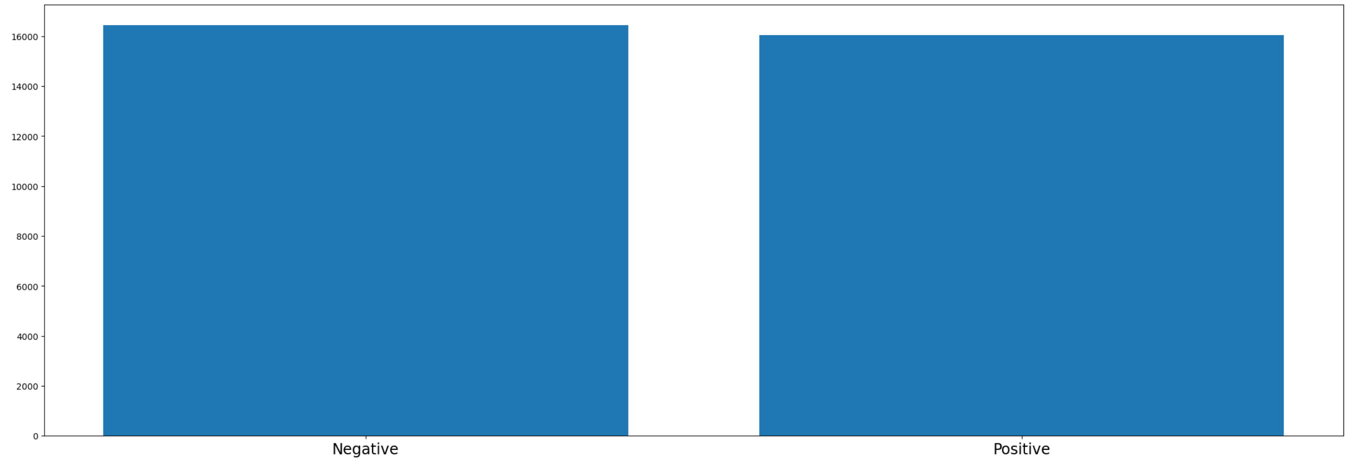


Figure 10: Feature distribution: dcmv.

#### dcmv

**Description:** Donor CMV test result.

**Type:** Categorical

- Negative : 0
- Positive : 1

**Density:** 1.0

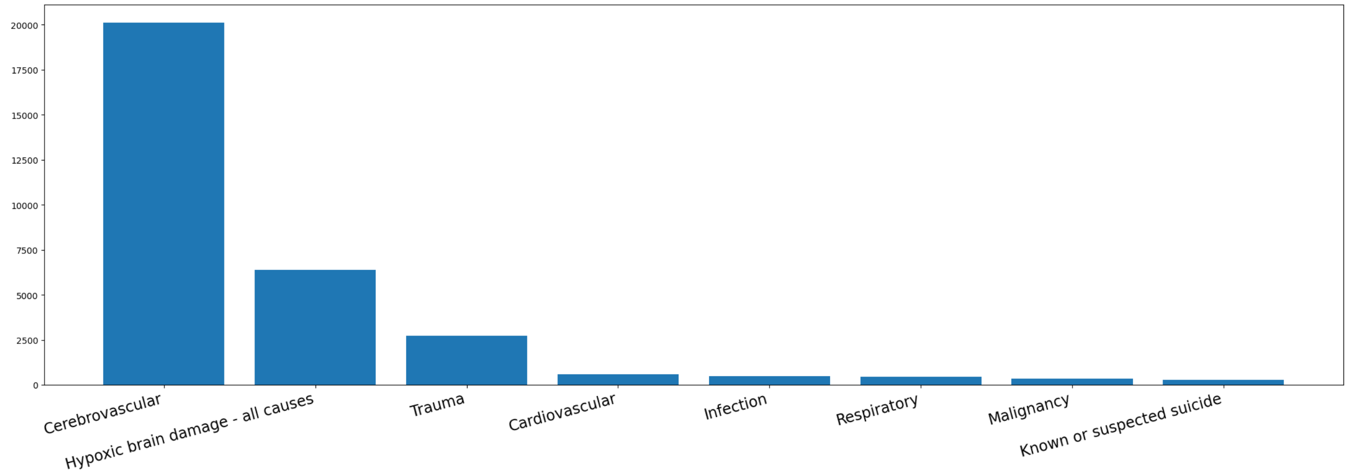


Figure 11: Feature distribution: dcod.

#### dcod

**Description:** Donor cause of death.

**Type:** Categorical

**Density:** 1.0

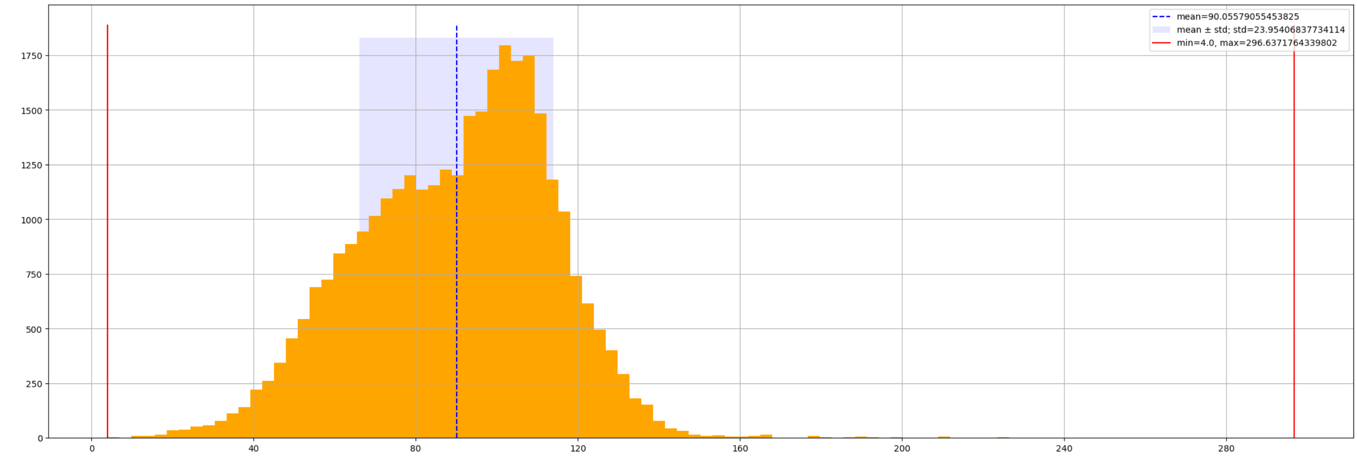


Figure 12: Feature distribution: degfr_base.

#### degfr_base

**Description:** Donor eGFR – first measure.

**Type:** Numerical

**Density:** 1.0

**Mininimum:** 4.0

**Maximum:** 296.6

**Mean:** 90.0

**Standard deviation:** 24.0

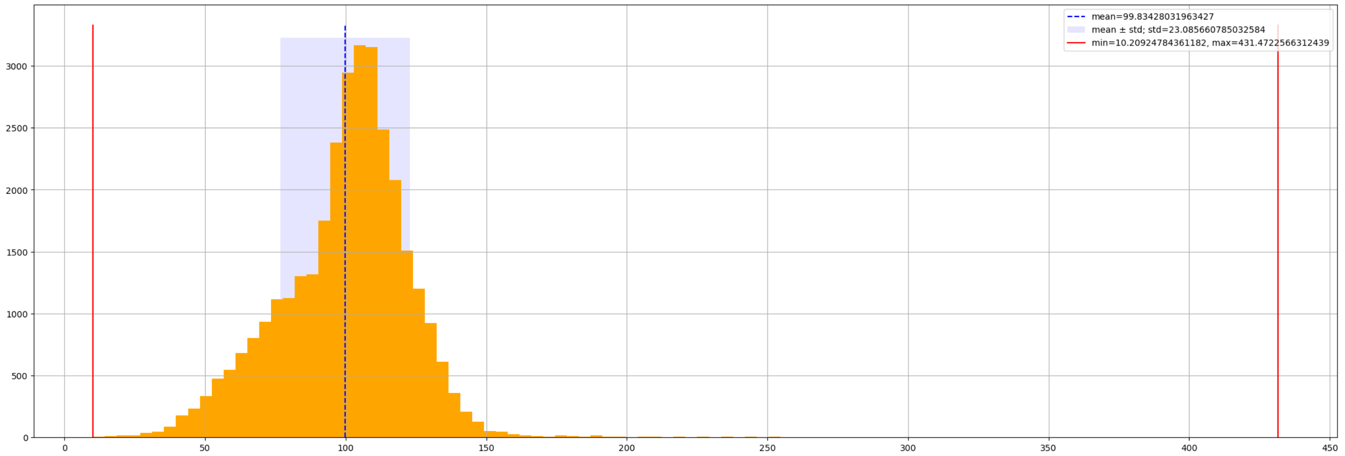


Figure 13: Feature distribution: degfr_max.

### degfr_max

**Description:** Maximum value for donor eGFR.

**Type:** Numerical

**Density:** 1.0

**Mininimum:** 10.2

**Maximum:** 431.5

**Mean:** 99.8

**Standard deviation:** 23.1

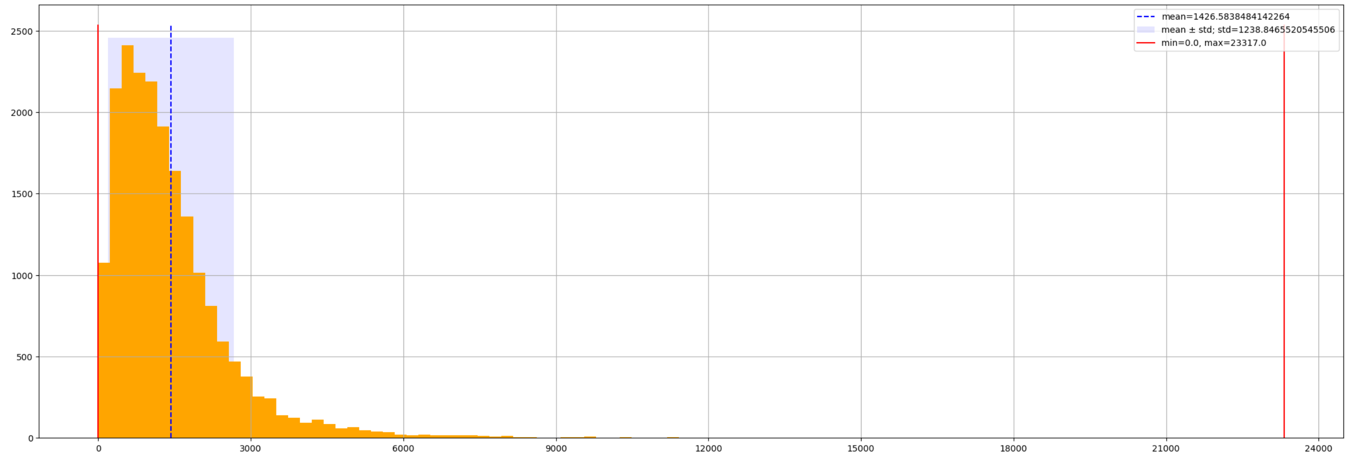

Figure 14: Feature distribution: dial_days.

#### dial_days

**Description:** Patient time on dialysis (days).

**Type:** Numerical

**Density:** 0.6

**Mininimum:** 0.0

**Maximum:** 23317.0

**Mean:** 1426.6

**Standard deviation:** 1238.9

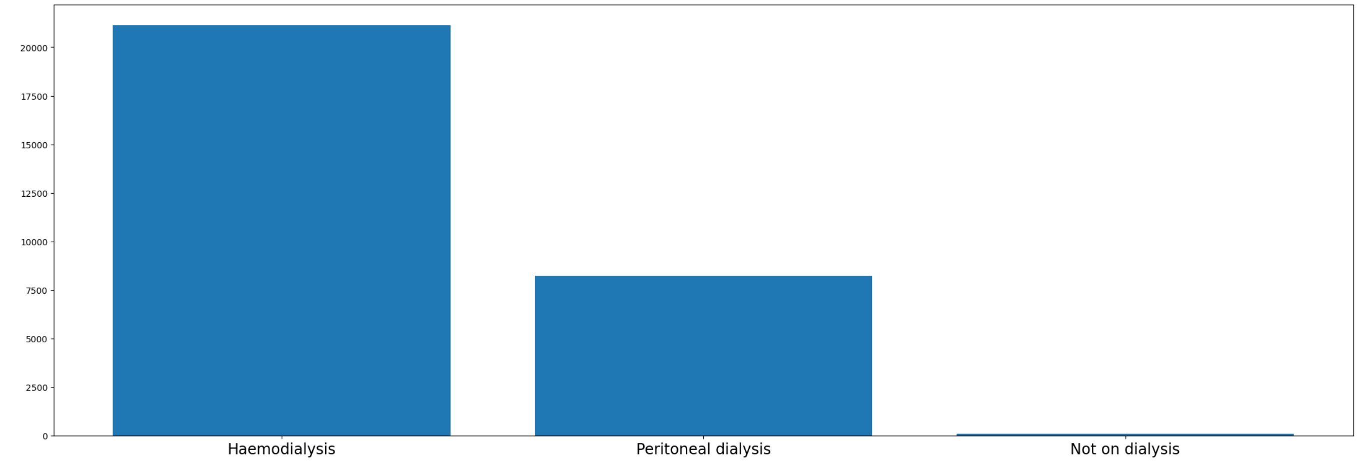

Figure 15: Feature distribution: dial_type.

### dial_type

**Description:** Most recent type of dialysis regarding transplantation.

**Type:** Categorical

**Density:** 0.9

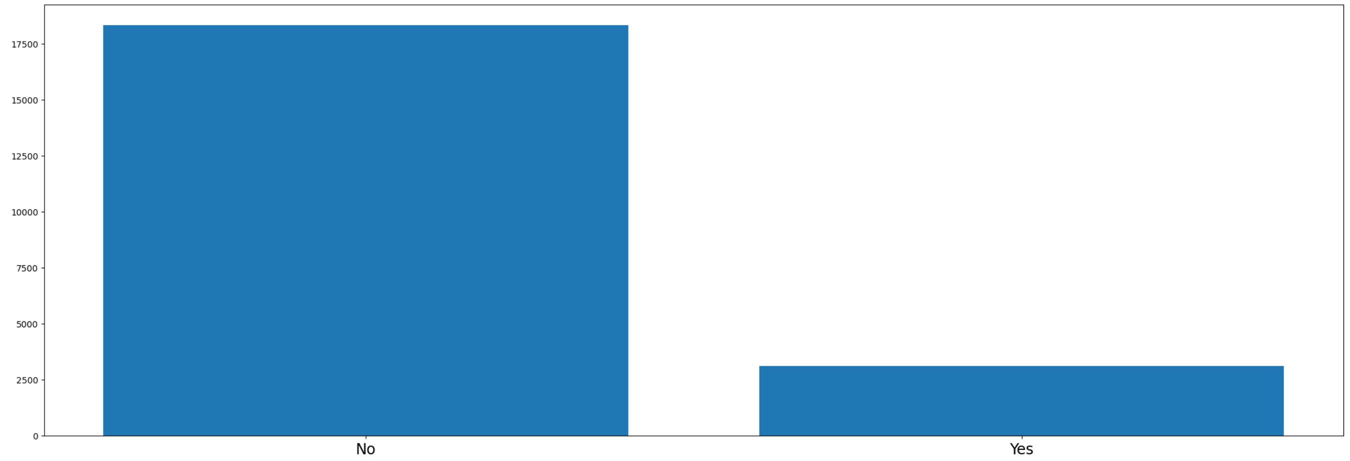


Figure 16: Feature distribution: dpast_alcohol_abuse.

**dpast_alcohol_abuse**

**Description:** Donor history of alcohol abuse.

**Type:** Categorical

- No : 0
- Yes : 1

**Density:** 0.7

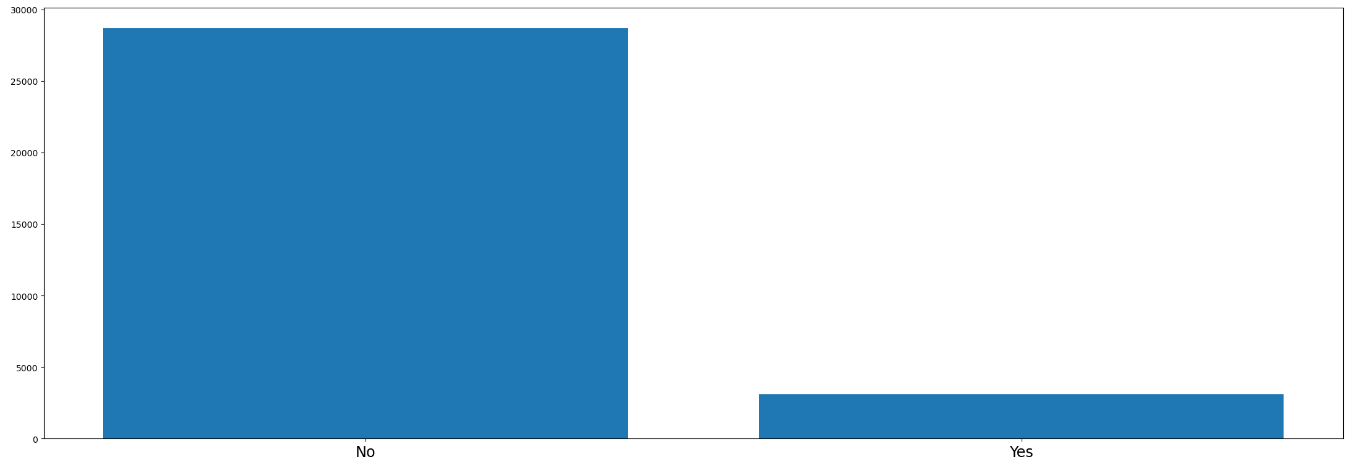


Figure 17: Feature distribution: dpast_cardio_disease.

**dpast_cardio_disease**

**Description:** Donor cardio disease.

**Type:** Categorical

- No : 0
- Yes : 1

**Density:** 1.0

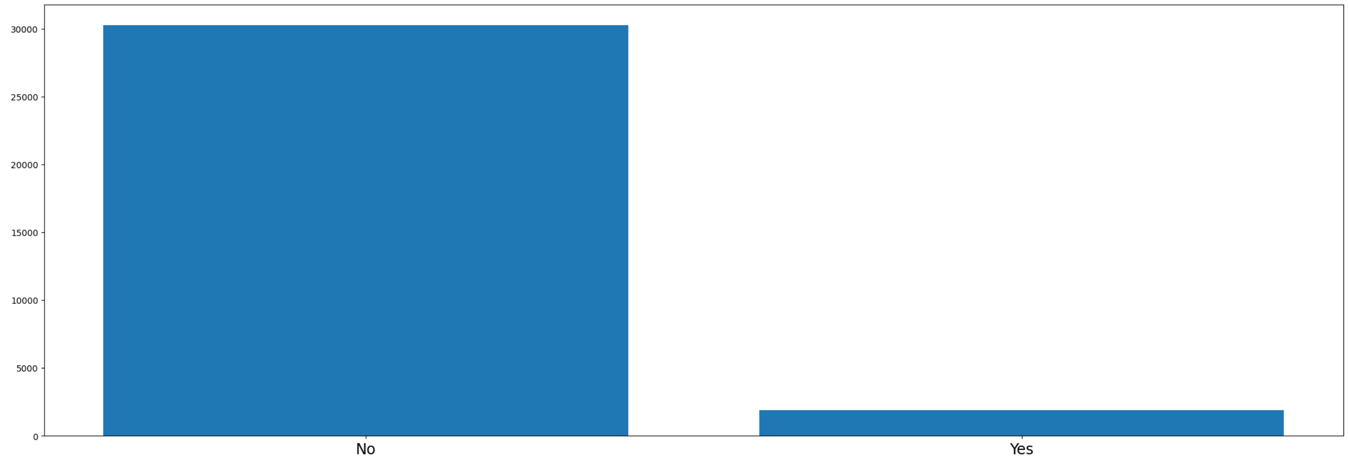


Figure 18: Feature distribution: dpast_diabetes.

**dpast_diabetes**

**Description:** Donor diabetes.

**Type:** Categorical

- No : 0
- Yes : 1

**Density:** 1.0

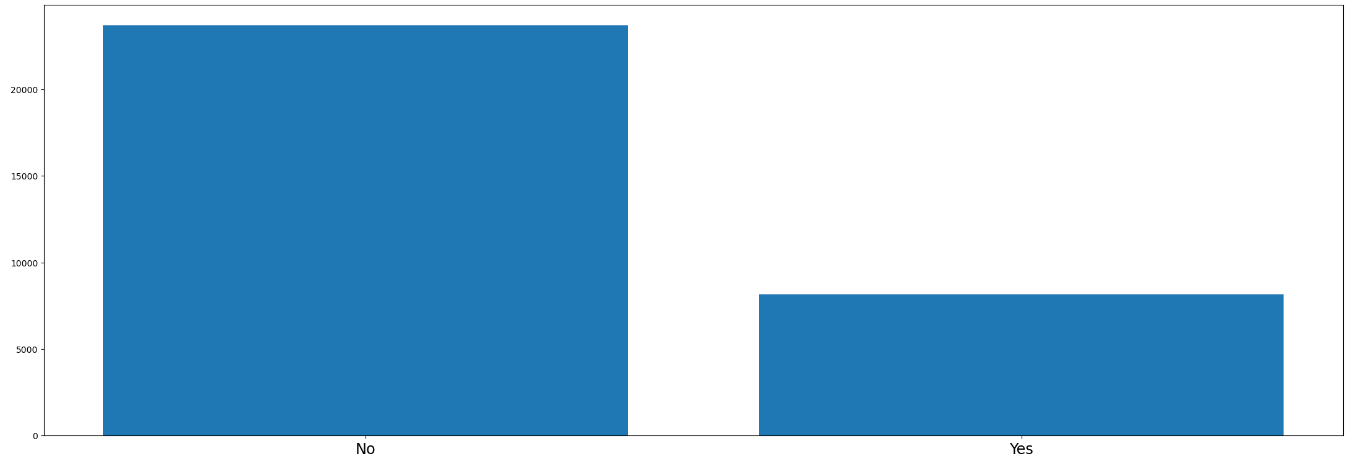


Figure 19: Feature distribution: dpast_hypertension.

#### dpast_hypertension

**Description:** Donor hypertension.

**Type:** Categorical

- No : 0
- Yes : 1

**Density:** 1.0

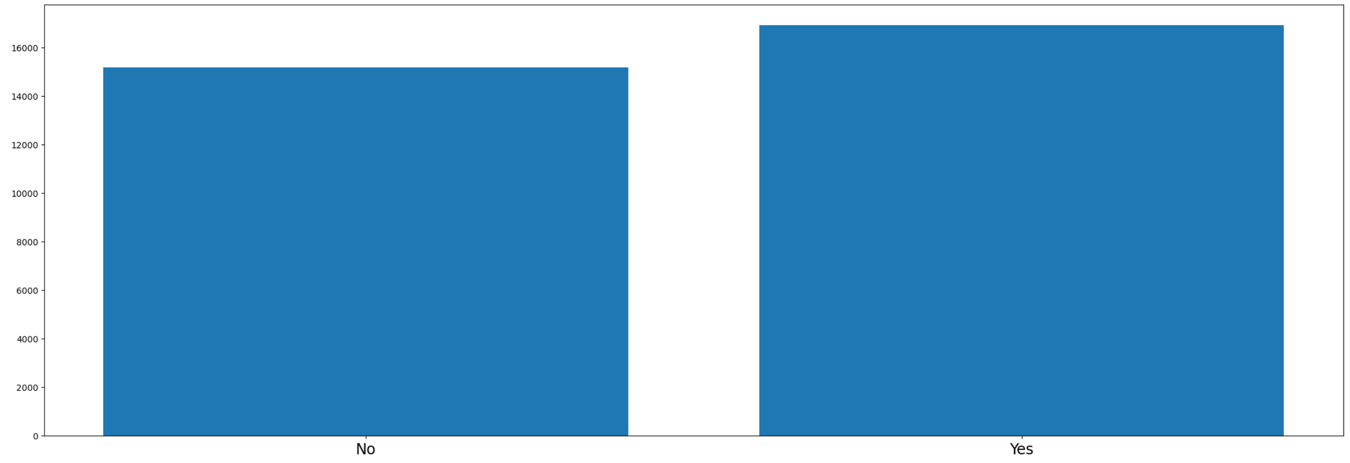


Figure 20: Feature distribution: dpast_smoker.

**dpast_smoker**

**Description:** Donor smoker.

**Type:** Categorical

- No : 0
- Yes : 1

**Density:** 1.0

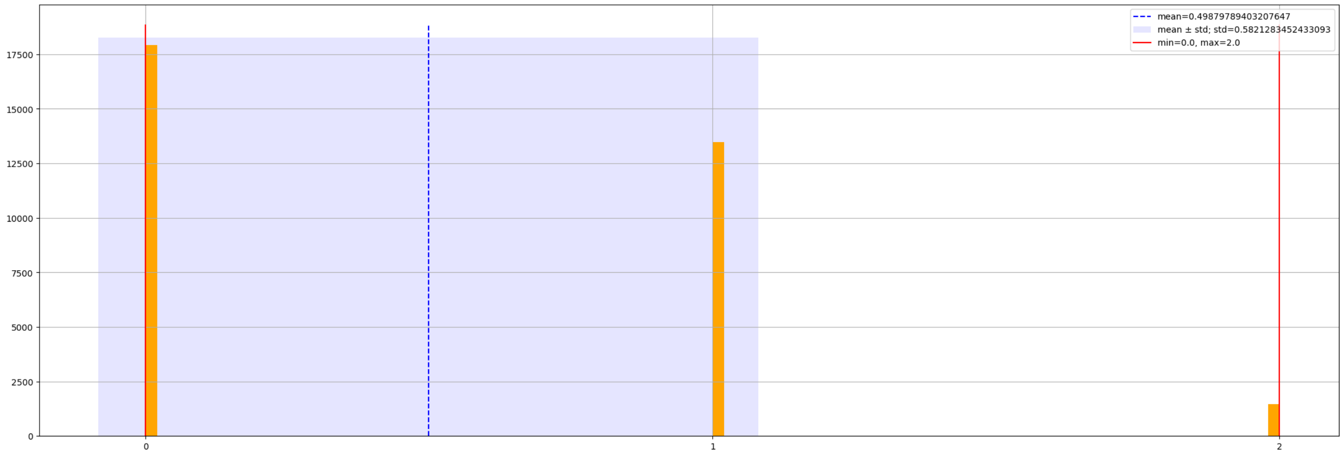


Figure 21: Feature distribution: drmm.

#### drmm

**Description:** Number of mismatches at DR locus.

**Type:** Numerical

**Density:** 1.0

**Mininimum:** 0.0

**Maximum:** 2.0

**Mean:** 0.5

**Standard deviation:** 0.6

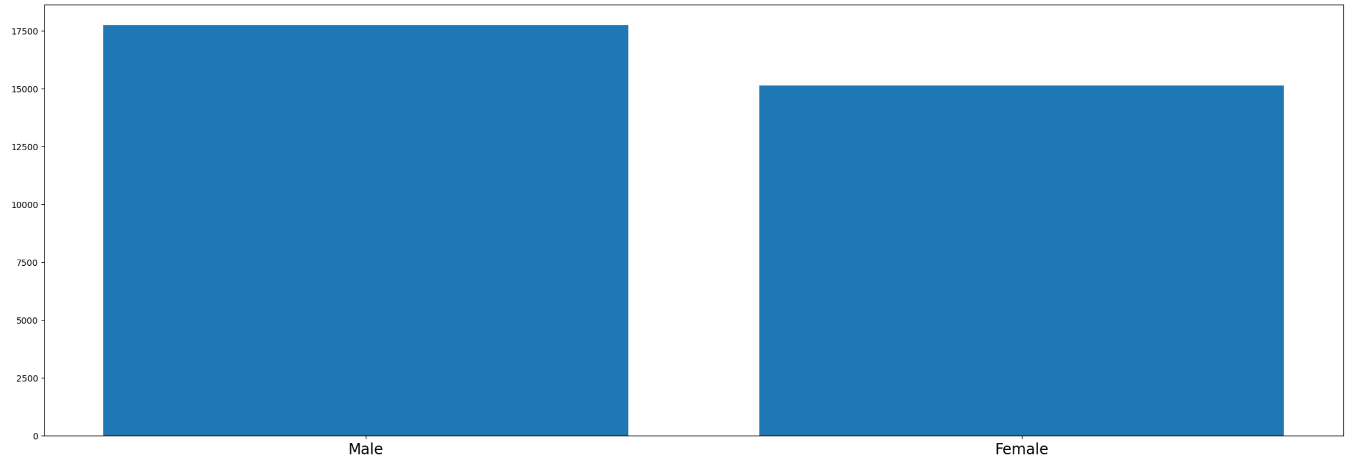


Figure 22: Feature distribution: dsex.

**dsex**

**Description:** Donor sex.

**Type:** Categorical

- Male : 0
- Female : 1

**Density:** 1.0

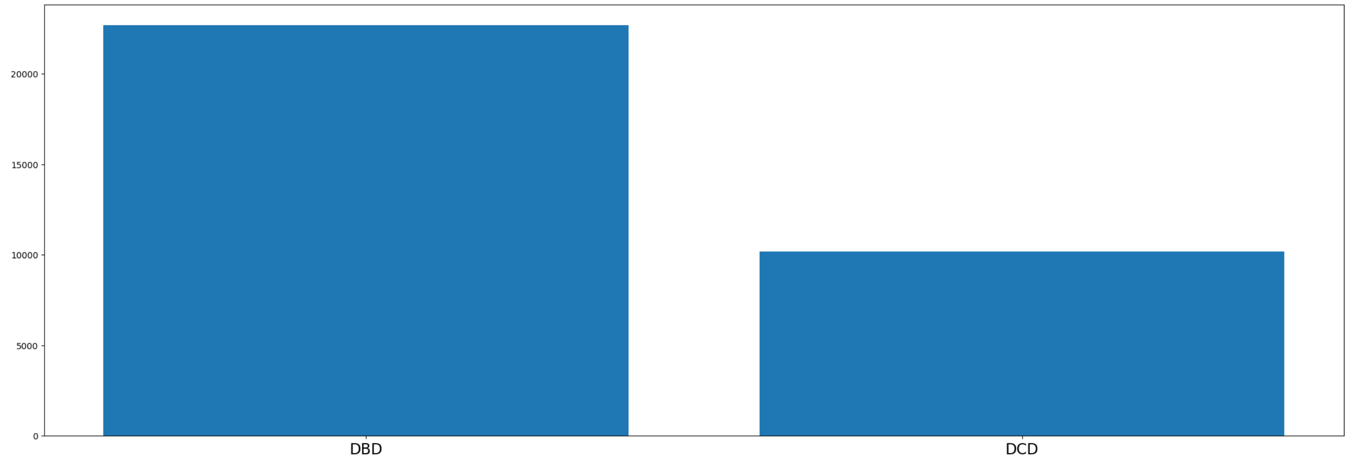


Figure 23: Feature distribution: dtype.

**dtype**

**Description:** Type of donor.

**Type:** Categorical

- DBD : 0
- DCD : 1

**Density:** 1.0

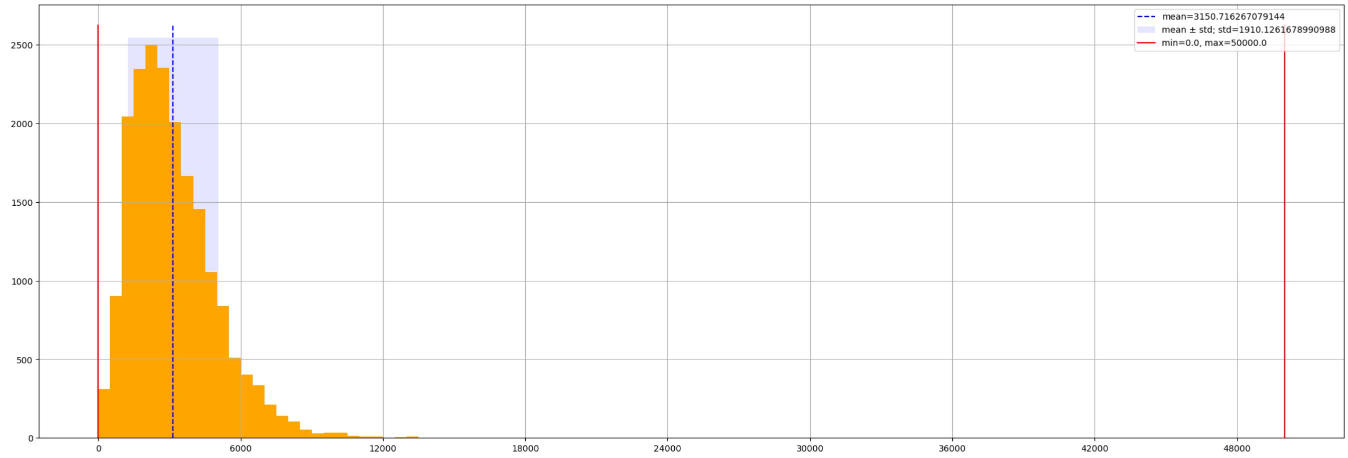


Figure 24: Feature distribution: durine_output_24hrs.

#### durine_output_24hrs

**Description:** Donor urine output within the last 24 hours (ml).

**Type:** Numerical

**Density:** 0.6

**Mininimum:** 0.0

**Maximum:** 50000.0

**Mean:** 3150.7

**Standard deviation:** 1910.1

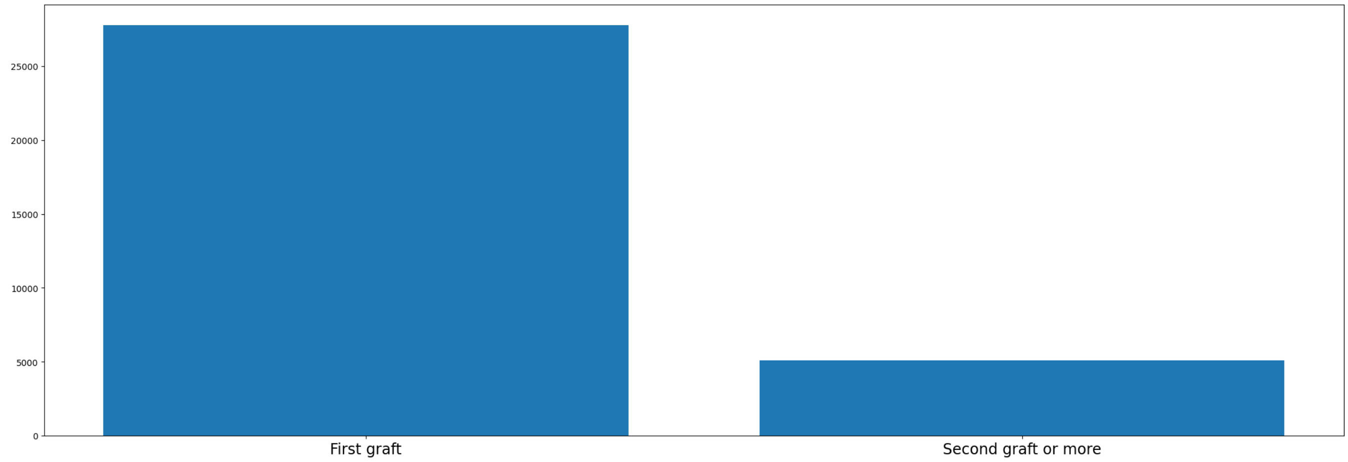


Figure 25: Feature distribution: graft_no.

**graft_no**

**Description:** Number of kidney transplants.

**Type:** Categorical

- First graft : 0
- Second graft or more : 1

**Density:** 1.0

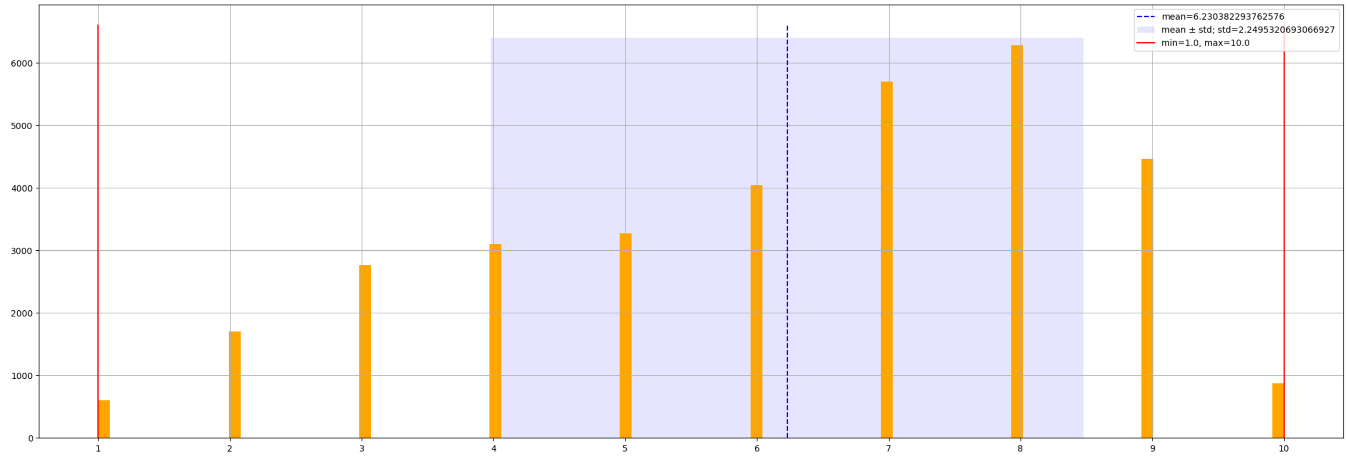


Figure 26: Feature distribution: matchbty.

#### matchbty

**Description:** Matchability.

**Type:** Numerical

**Density:** 1.0

**Mininimum:** 1.0

**Maximum:** 10.0

**Mean:** 6.2

**Standard deviation:** 2.3

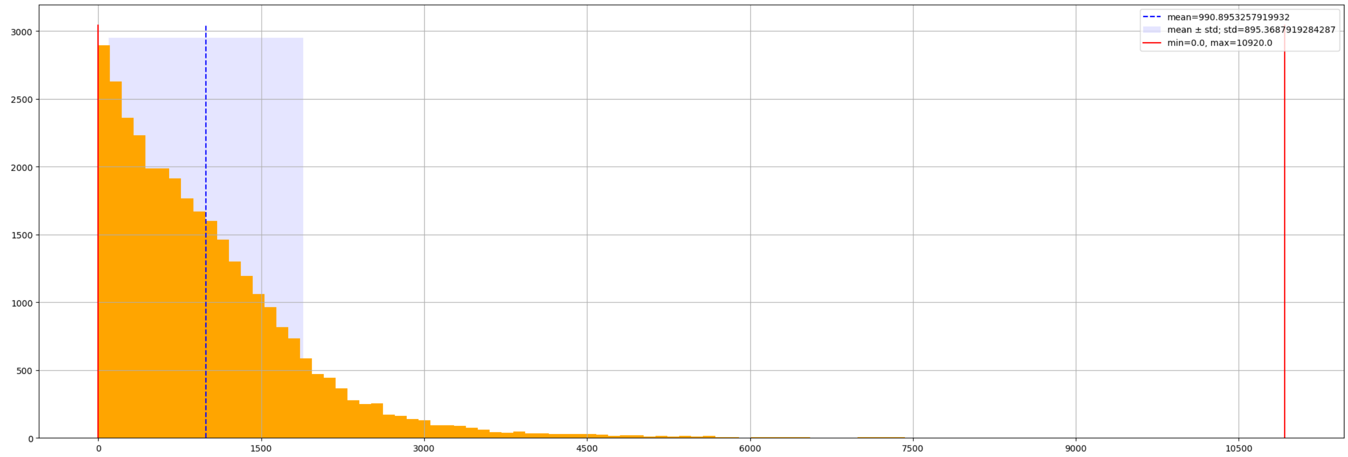


Figure 27: Feature distribution: offer_wait.

#### offer_wait

**Description:** Recipient waiting time (days), from registration to offer.

**Type:** Numerical

**Density:** 1.0

**Mininimum:** 0.0

**Maximum:** 10920.0

**Mean:** 990.9

**Standard deviation:** 895.4

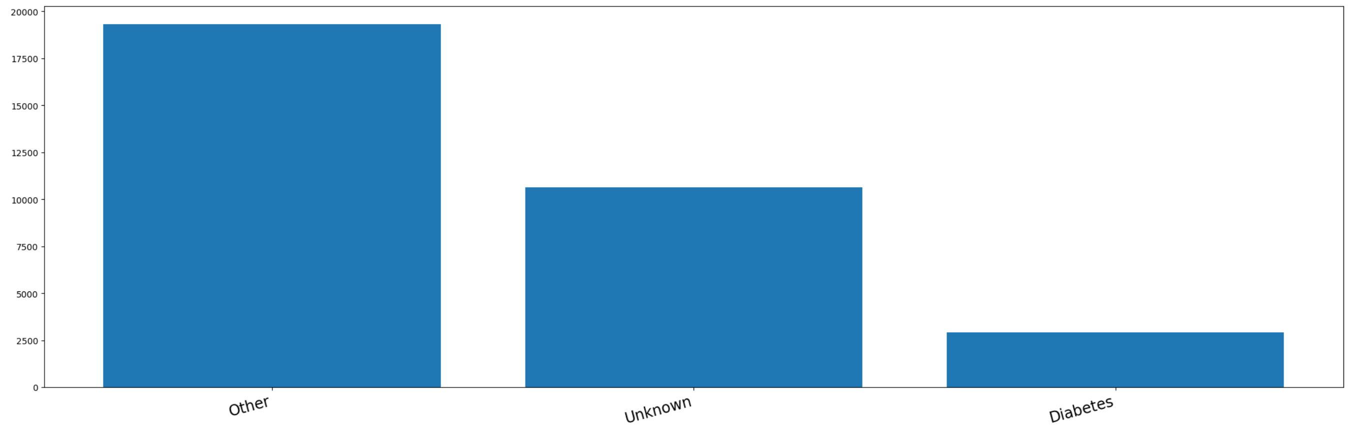


Figure 28: Feature distribution: prd.

#### prd

**Description:** Primary renal disease.

**Type:** Categorical

**Density:** 1.0

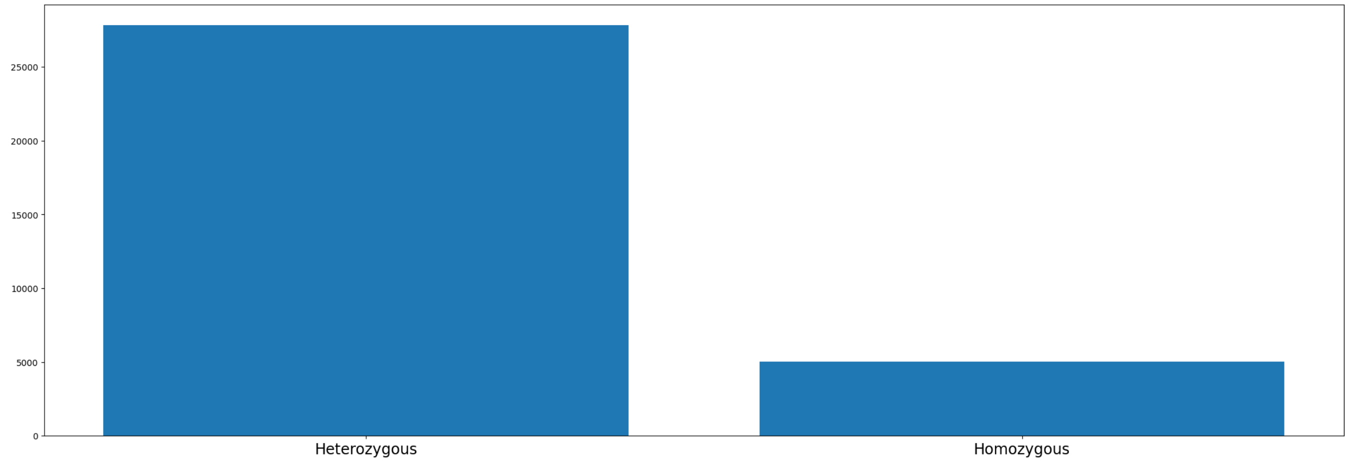


Figure 29: Feature distribution: r_dr_hom.

#### r_dr_hom

**Description:** Recipient homozygous at DR locus.

**Type:** Categorical

- Heterozygous : 0
- Homozygous : 1

**Density:** 1.0

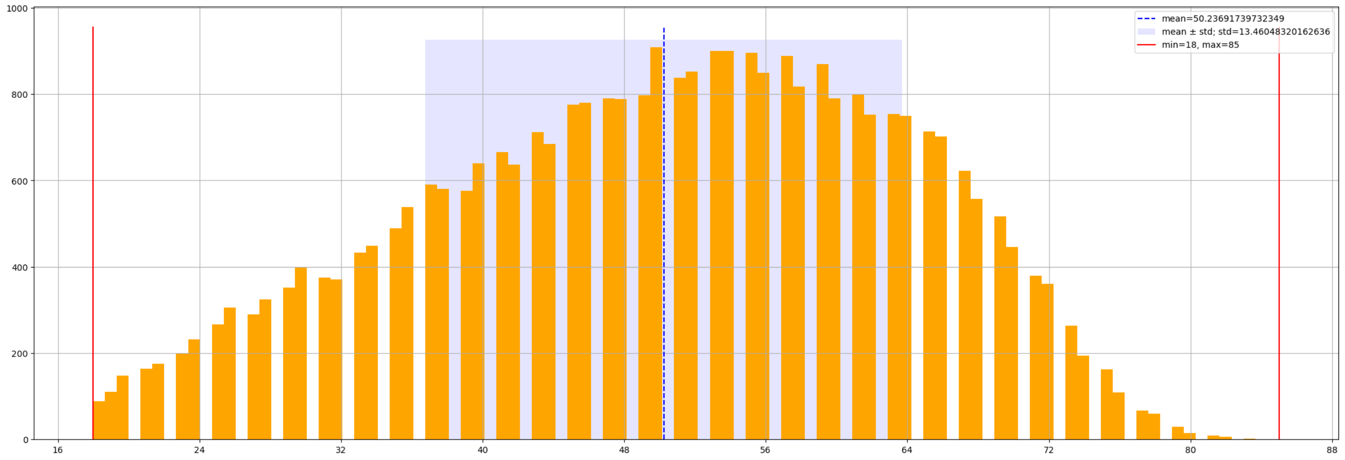


Figure 30: Feature distribution: rage.

**rage**

**Description:** Recipient age.

**Type:** Numerical

**Density:** 1.0

**Mininimum:** 18.0

**Maximum:** 85.0

**Mean:** 50.2

**Standard deviation:** 13.5

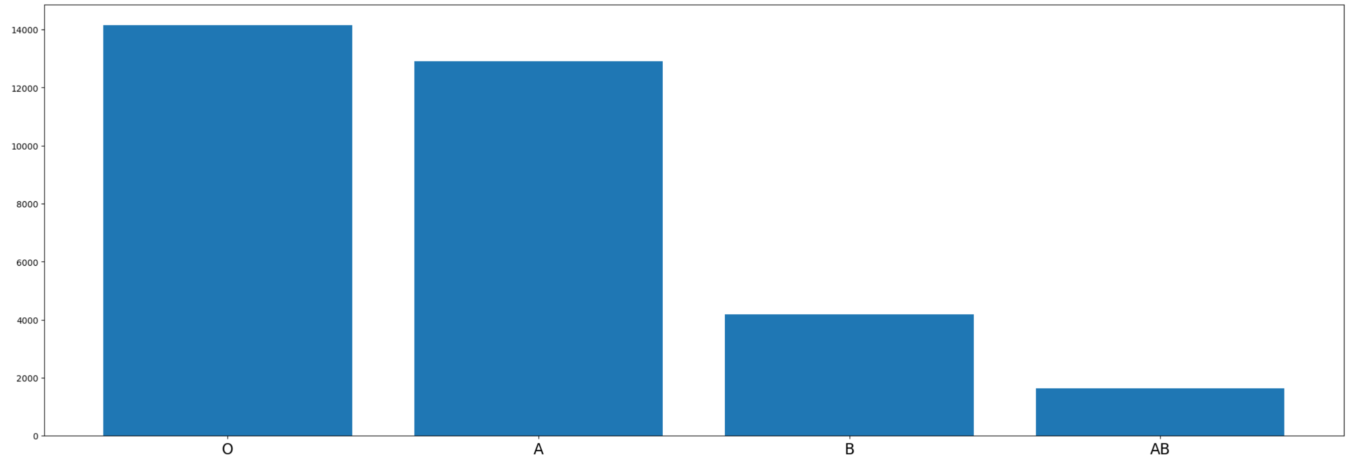


Figure 31: Feature distribution: rbg.

#### rbg

**Description:** Recipient blood group.

**Type:** Categorical

**Density:** 1.0

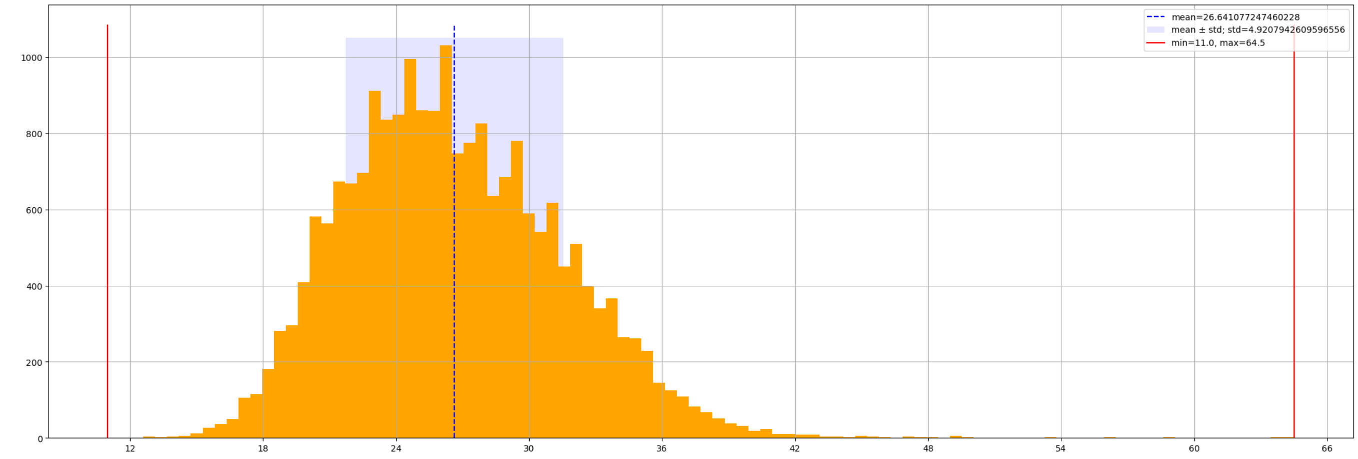


Figure 32: Feature distribution: rbmi.

#### rbmi

**Description:** Recipient BMI.

**Type:** Numerical

**Density:** 0.6

**Mininimum:** 11.0

**Maximum:** 64.5

**Mean:** 26.6

**Standard deviation:** 4.9

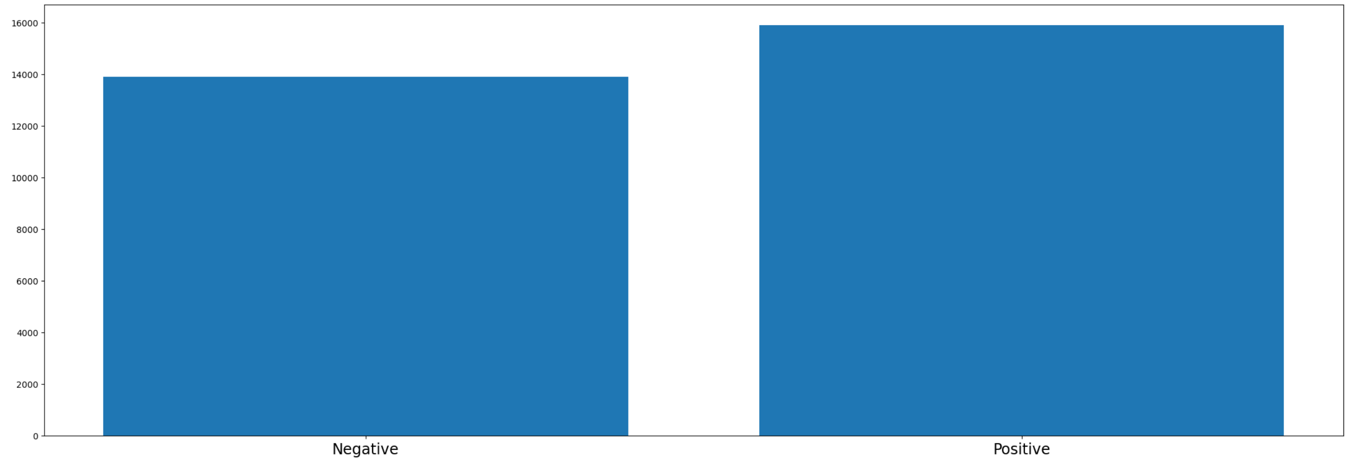


Figure 33: Feature distribution: rcmv.

**rcmv**

**Description:** Recipient CMV test result.

**Type:** Categorical

- Negative : 0
- Positive : 1

**Density:** 0.9

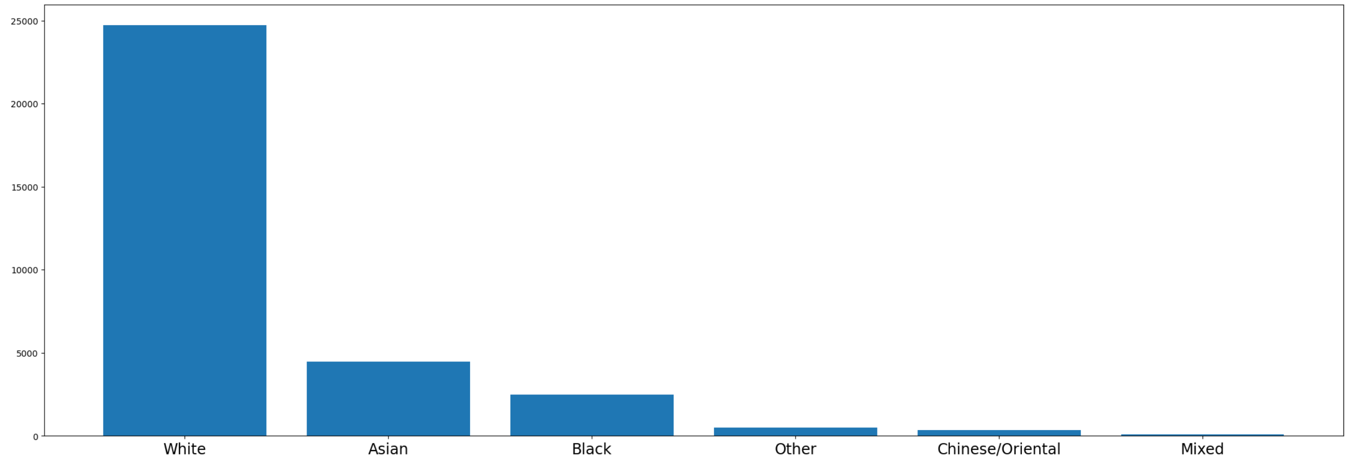


Figure 34: Feature distribution: rethnic.

**rethnic**

**Description:** Recipient ethnicity.

**Type:** Categorical

**Density:** 1.0

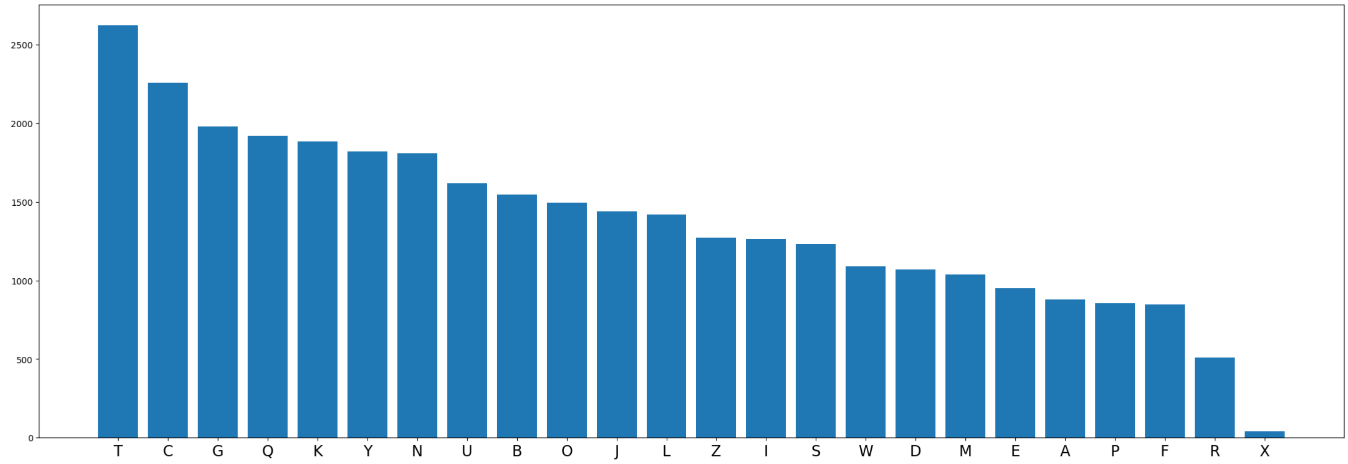


Figure 35: Feature distribution: rhosp.

#### rhosp

**Description:** Anonymised unit that received and transplanted the organ.

**Type:** Categorical

**Density:** 1.0

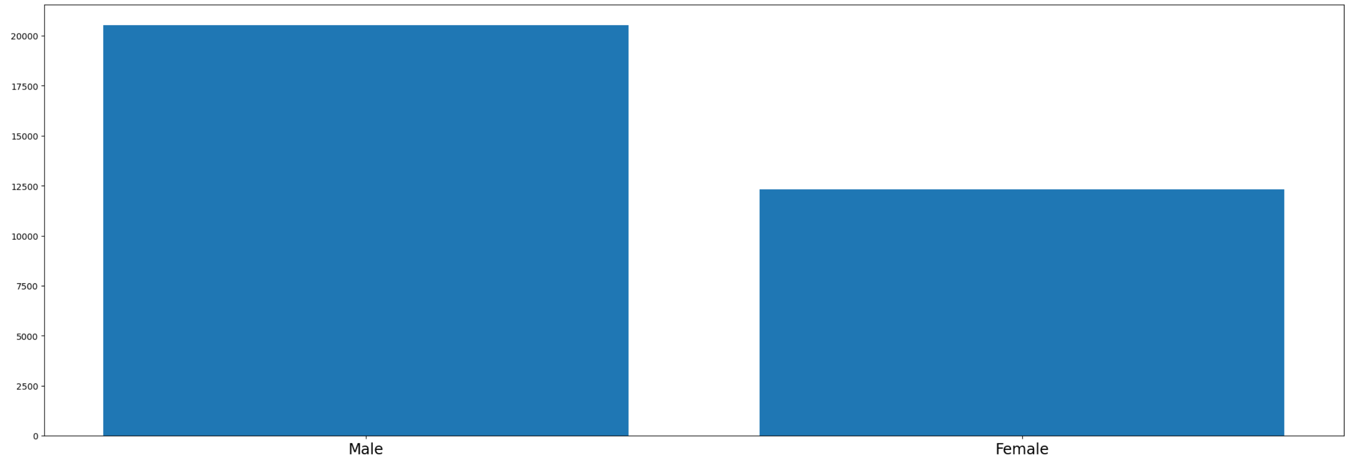


Figure 36: Feature distribution: rsex.

**rsex**

**Description:** Recipient sex.

**Type:** Categorical

- Male : 0
- Female : 1

**Density:** 1.0

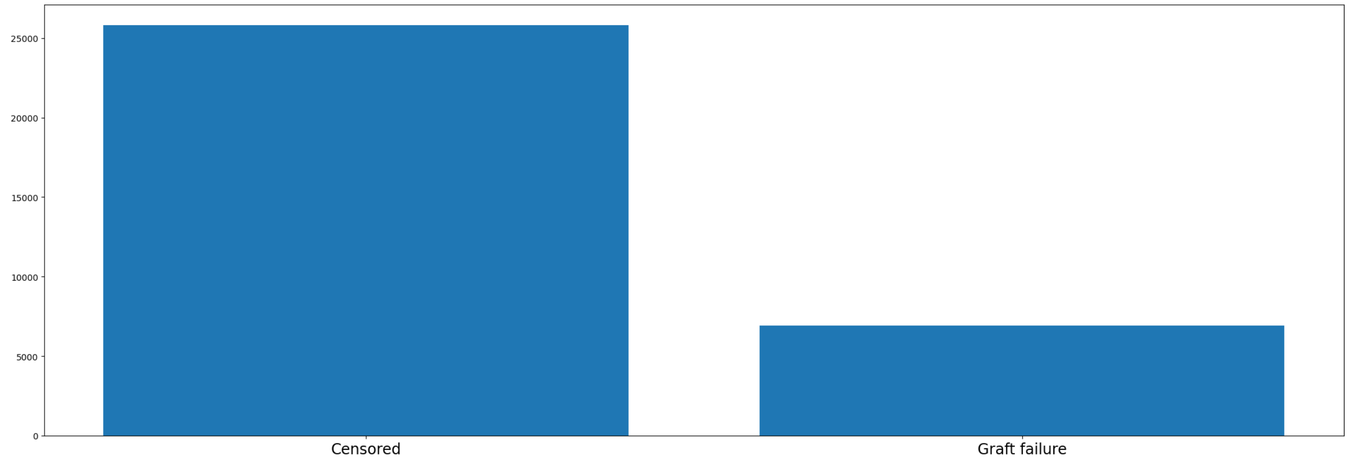


Figure 37: Target distribution: gcens.

**gcens**

**Description:** Graft censoring indicator.

**Type:** Categorical

- Censored : 0
- Graft failure : 1

**Density:** 1.0

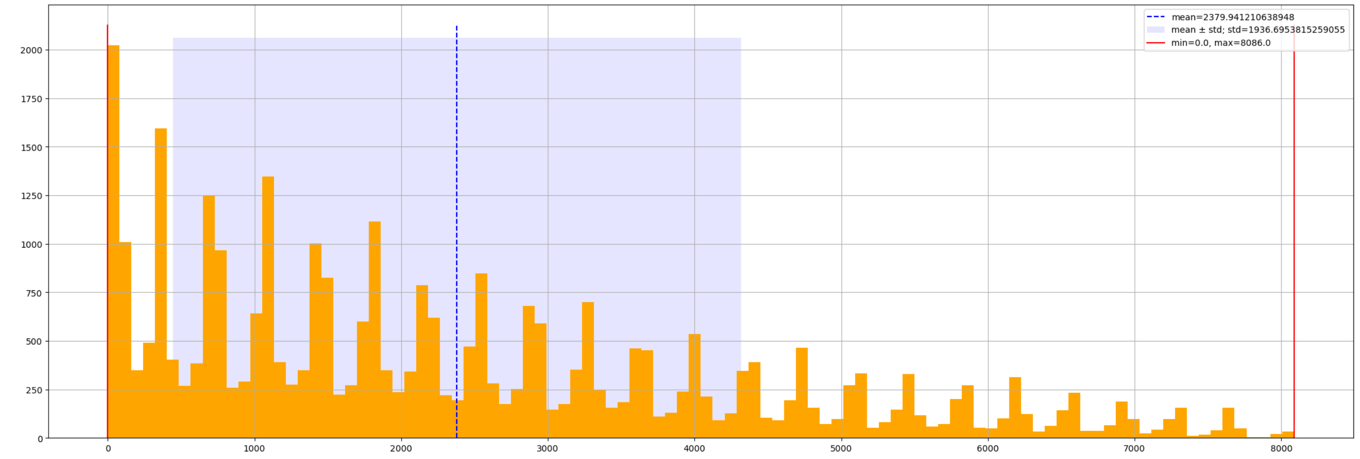


Figure 38: Target distribution: gsurv.

#### gsurv

**Description:** Graft survival time (days).

**Type:** Numerical

**Density:** 1.0

**Mininimum:** 0.0

**Maximum:** 8086.0

**Mean:** 2379.9

**Standard deviation:** 1936.7

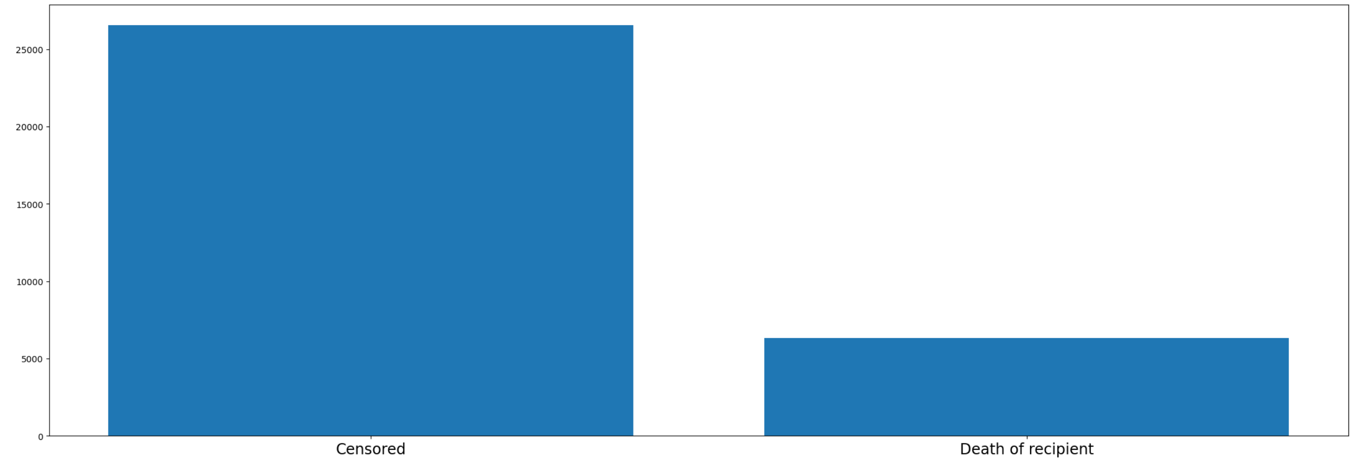


Figure 39: Target distribution: pcens.

**pcens**

**Description:** Patient censoring indicator.

**Type:** Categorical

- Censored : 0
- Patient death : 1

**Density:** 1.0

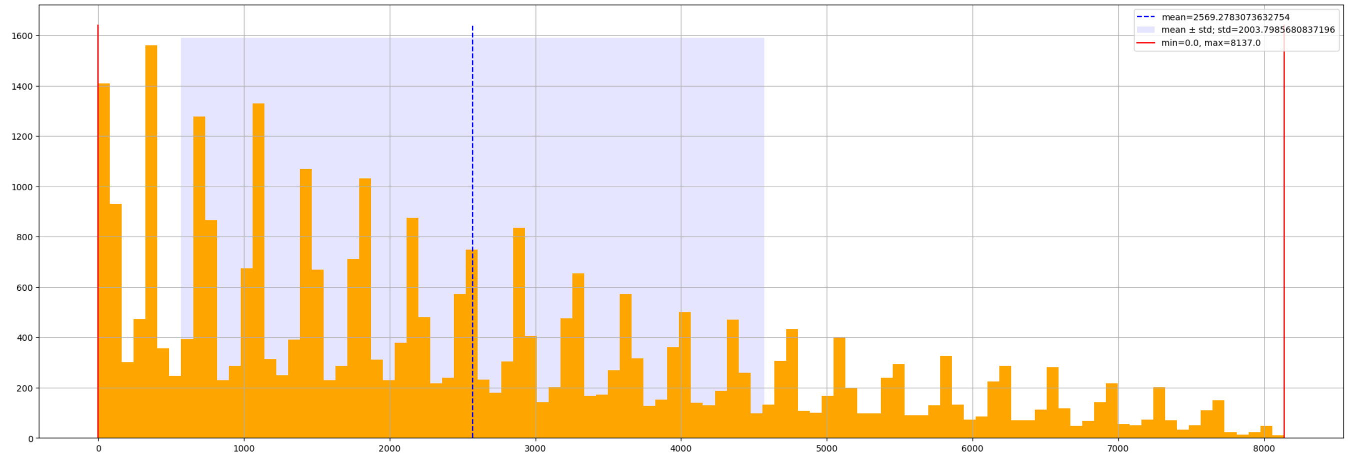


Figure 40: Target distribution: psurv.

#### psurv

**Description:** Patient survival time (days).

**Type:** Numerical

**Density:** 1.0

**Mininimum:** 0.0

**Maximum:** 8137.0

**Mean:** 2569.3

**Standard deviation:** 2003.8
